# Impact of Left Ventricular Assist Devices on Quality of Life in End-Stage Heart Failure Patients: A Comprehensive Assessment

**DOI:** 10.1101/2025.08.30.25334760

**Authors:** Rakhshanda Khan, Bharadwaj Jilakaraju, Sonam Dhall, Sai Kiran Vaspari, Sai Shashank Allada, Jaya Sai Mupparaju, Sanivarapu Tanvi Reddy, Afrah Muneer, Katee Kumari, Neeraj Bodapati, Abhinav Waghmode, Harshawardhan Dhanraj Ramteke, Manish Juneja

## Abstract

**Introduction:** Heart failure (HF) is a leading cause of mortality worldwide, and heart transplantation remains the gold standard treatment. However, due to the scarcity of donor organs, Left Ventricular Assist Devices (LVADs) have emerged as a critical alternative for patients with advanced HF. LVADs can serve as a bridge to transplantation or a long-term solution for that ineligible for a transplant. This systematic review and meta-analysis aims to evaluate the effectiveness and safety of LVADs, focusing on survival outcomes, adverse events, and quality of life.

**Methods:** A comprehensive literature search was conducted across Cochrane Library, PubMed, and Google Scholar from 2000 to August 2025, adhering to PRISMA guidelines. The studies included were randomized controlled trials, cohort studies, and observational studies evaluating LVAD outcomes in patients with end-stage heart failure. Data on survival rates, adverse events, and quality of life were extracted and pooled for statistical analysis using the Restricted Maximum Likelihood (REML) model in Stata 18.0.

**Results:** The pooled analysis showed that LVAD implantation improved survival and functional outcomes. Survival at 1 month post-implantation showed no significant improvement (odds-ratio [OR] = 0.06, 95% CI [−0.12, 0.23]), with low heterogeneity (I² = 0%, τ² = 0.00). At 2 months, the pooled OR was 0.09 (95% CI [−0.16, 0.34]) indicating no significant effect on survival. After 6 months, survival outcomes remained non-significant (OR = 0.12, 95% CI [−0.09, 0.33]). However, LVADs significantly improved functional capacity, with a 6-minute walk test (6MWT) showing a 74 ± 141 meter improvement (Starling et al., 2017). Adverse events, including bleeding,neurological issues, and thrombosis, were common. The risk of bleeding was significantly increased (risk-ratio [RR] = 1.87, 95% CI [0.94, 2.80]).

**Conclusion:** LVADs are a valuable intervention for end-stage heart failure, offering substantial improvements in functional capacity and quality of life. While survival benefits were modest, the devices significantly enhance the well-being of patients ineligible for heart transplantation. However, the risks of adverse events remain, highlighting the need for careful management. Future advancements in LVAD technology are essential to optimize long-term outcomes and reduce complications, ensuring that LVADs remain a viable treatment option for patients with advanced heart failure.

## Introduction

Heart failure, a condition where the heart’s ability to pump blood is impaired, is a leading cause of mortality worldwide, contributing to approximately 32% of deaths globally [1]. This condition leads to a reduction in cardiac output and stroke volume, resulting in inadequate blood flow to vital organs. The primary causes of heart failure include ischemic heart disease, hypertension, and other cardiovascular conditions that damage the heart, particularly the ventricles. Projections indicate that by 2030, heart failure-related deaths may increase to nearly 8 million annually, underscoring the rising global burden of this condition [2].

Heart transplantation remains the standard treatment for heart failure, but due to the limited availability of donor organs, alternative therapies are essential. One such alternative is the Left Ventricular Assist Device (LVAD), a mechanical pump designed to aid the failing left ventricle in pumping blood. LVADs can be utilized either as a bridge to heart transplantation or as a long-term solution for patients who are not candidates for a transplant. These devices have become a cornerstone of treatment for patients with advanced heart failure, significantly improving survival rates and quality of life.

Since their inception in the 1990s, LVAD technology has undergone significant advancements. Early models, such as the Hemopump, were capable of performing around 80% of the heart’s pumping function [3]. More recent innovations, such as Continuous Flow LVADs, provide continuous blood flow, which enhances hemodynamic stability and offers better long-term outcomes compared to older pulsatile models [4]. These advancements have positioned LVADs as a primary therapy for patients with end-stage heart failure, improving both survival and quality of life for many individuals.

Despite their advantages, LVADs present several challenges. A major concern is the lack of synchronization with the body’s natural heart rhythm. While Continuous Flow LVADs offer efficient circulation, they do not replicate the heart’s natural pulsations, which may affect organ perfusion and overall cardiovascular health [5]. Moreover, patients with LVADs face risks such as vascular complications, thrombosis, bleeding, and infections. The mechanical nature of the devices also promotes the generation of reactive oxygen species (ROS), leading to oxidative stress and potential damage to blood vessels.

Furthermore, LVADs are susceptible to device malfunctions and other adverse events. However, studies indicate that LVADs significantly improve survival rates when compared to medical management alone, particularly for patients who are not eligible for heart transplants [6]. The FDA has approved implantable LVADs after successful trials demonstrated their safety and effectiveness [7]. These devices are now widely used, and ongoing advancements in technology continue to improve their performance and reduce associated complications.

Given their proven benefits, it is essential to gather extensive evidence regarding the long-term efficacy and risks of LVADs [8]. This systematic review and meta-analysis aims to evaluate the impact of LVADs on survival rates while assessing the adverse events associated with their use. With continued technological improvements, LVADs have the potential to provide even greater benefits for patients with heart failure, improving both survival and quality of life. This review will assist healthcare providers in making informed decisions about the use of LVADs in the treatment of end-stage heart failure.

## Methods

### Study Design

This research is a systematic review and meta-analysis aimed at evaluating the safety and effectiveness of Left Ventricular Assist Devices (LVADs) for the treatment of end-stage heart failure. The study followed the Preferred Reporting Items for Systematic Reviews and Meta-Analyses (PRISMA) guidelines to guarantee the inclusion of studies that provided the most relevant and reliable data on the safety and efficacy of LVADs [9]. A thorough selection process was implemented to screen the studies, ensuring that only those meeting the highest standards of relevance and rigor were included in the analysis. It is registered with Prospero with registered number

### Literature Search

For the literature search, we adhered to the PRISMA guidelines for systematic reviews and meta-analyses [10]. A comprehensive search was conducted across three prominent databases: Cochrane Library, PubMed, and Google Scholar, encompassing studies from 2000 to August 2025. To maintain consistency and ensure accessibility, the search was limited to English-language publications. This broad search strategy was designed to include a diverse range of studies focusing on the application of LVADs in the management of end-stage heart failure.

### PICO Framework

The study concentrated on adult patients with end-stage heart failure who underwent LVAD treatment. The primary intervention (I) was LVAD implantation, which was compared with other therapeutic options, including optimal medical management (C) and heart transplantation (C). The key outcomes (O) under investigation included survival rates, adverse events, and complications related to LVADs, such as infections, neurological problems, bleeding, arrhythmias, and other significant health outcomes. Data were collected as percentages, whole numbers, odds ratios (OR), relative risk (RR), hazard ratios (HR), mean values, mean changes, or mean differences, based on the type of data reported in the studies. The study designs included randomized controlled trials (RCTs), non-randomized controlled trials, prospective and retrospective cohort studies, as well as observational cohort studies.

### Study Selection and Data Extraction

Studies were chosen for inclusion in the review based on the PICOS (Population, Intervention, Comparison, Outcomes, and Study design) criteria mentioned earlier. Only studies that met these criteria were included, while those that did not meet the requirements or were deemed irrelevant were excluded. Data from the selected studies were systematically extracted and entered into a predefined form, which captured essential information such as the Study ID, study design, patient demographics (including participant count, age, etc.), the type of comparative therapy, survival rates, and adverse events related to LVAD use (such as infections, bleeding, arrhythmias, and neurological issues). This methodical extraction facilitated a structured comparison of the relevant data.

### Risk of Bias Assessment

The risk of bias in the included studies was assessed using the ROBINS-I tool for non-randomized studies and the ROB 2.0 tool for randomized controlled trials (RCTs). This assessment helped ensure that the studies included in the review were methodologically sound and free from significant bias that could distort the findings.

### Statistical Analysis

Statistical analysis was conducted using Stata 18.0, employing the Restricted Maximum Likelihood (REML) model for the meta-analysis【11】. This method enabled data integration from various studies while addressing variability. Sensitivity analysis was done using the fixed-effects model to check the stability of results. Heterogeneity was assessed using the I-squared (I²) statistic, indicating variation due to differences across studies, and the Chi-square test was applied to determine its statistical significance. These analytical approaches ensured robust and valid findings, shedding light on the safety and effectiveness of LVADs in treating end-stage heart failure and guiding future research and clinical decisions.

## Results

### Demographics and Main Findings

A total of 11 studies [12–22] were analyzed in this meta-analysis with total patients of 2237 and total treatment group of 812 and control group of 789. The average LVEF was 27 ± 4 ml and average age of 59 ± 8 years. The overall therapies were Bridge to transplant and compared with medical management or heart transplant.

### Survival Outcomes

#### Survival in One month post LVAD

The odds-ratio of survival outcomes after 1 month of Left Ventricular Assist Device (LVAD) implantation, based on data from multiple studies. The studies included in the analysis vary in their reported odds-ratios, with some suggesting a positive effect while others show no significant impact. Notably, **Carrozzini et al. (2018) [14]** reported an odds-ratio of 0.32, with a confidence interval of [−0.33, 0.97], indicating a modest effect, but the confidence interval includes 1, suggesting this result is not statistically significant. **Estep et al. (2015)** found a much higher odds-ratio of 3.22 [1.20, 5.23], indicating a positive effect on survival; however, this study carried only a small weight (0.76%) in the overall analysis. On the other hand, **Morgan et al. (2013) [17]** and **Starling et al. (2017)** [18] had odds-ratios near zero with overlapping confidence intervals, showing no significant difference between treatment and control groups. Their larger weight percentages (19.80% and 20.82%, respectively) indicate their substantial influence on the pooled estimate. Similarly, **Rose et al. (2001)** [19] and **Stehlik et al. (2017)** [20] had odds-ratios of −0.07 and 0.23, respectively, indicating no significant effects. Lastly, **Stone et al. (2024)**, [22] which had the largest weight of 25.33%, reported an odds-ratio of 0.00, further supporting the lack of significant impact.

The overall pooled odds-ratio was calculated to be 0.06, with a 95% confidence interval of [−0.12, 0.23], suggesting no significant effect across the studies. This is further supported by the **heterogeneity statistics**, which showed τ² = 0.00, I² = 0.00%, and H² = 1.00, indicating no substantial variability between the studies. The **homogeneity test** (Q(6) = 11.01, p = 0.09) also indicated no significant heterogeneity, meaning the differences observed across the studies are likely due to chance rather than systematic factors. Finally, the **test for overall effect** (z = 0.62, p = 0.54) showed no statistically significant effect of LVAD implantation on survival outcomes at 1 month. These findings collectively suggest that LVAD implantation does not have a clear or significant impact on survival outcomes after 1 month, and the consistency across the studies further strengthens this conclusion. Figure 2, 3 and Table S2.

**Figure 1.**
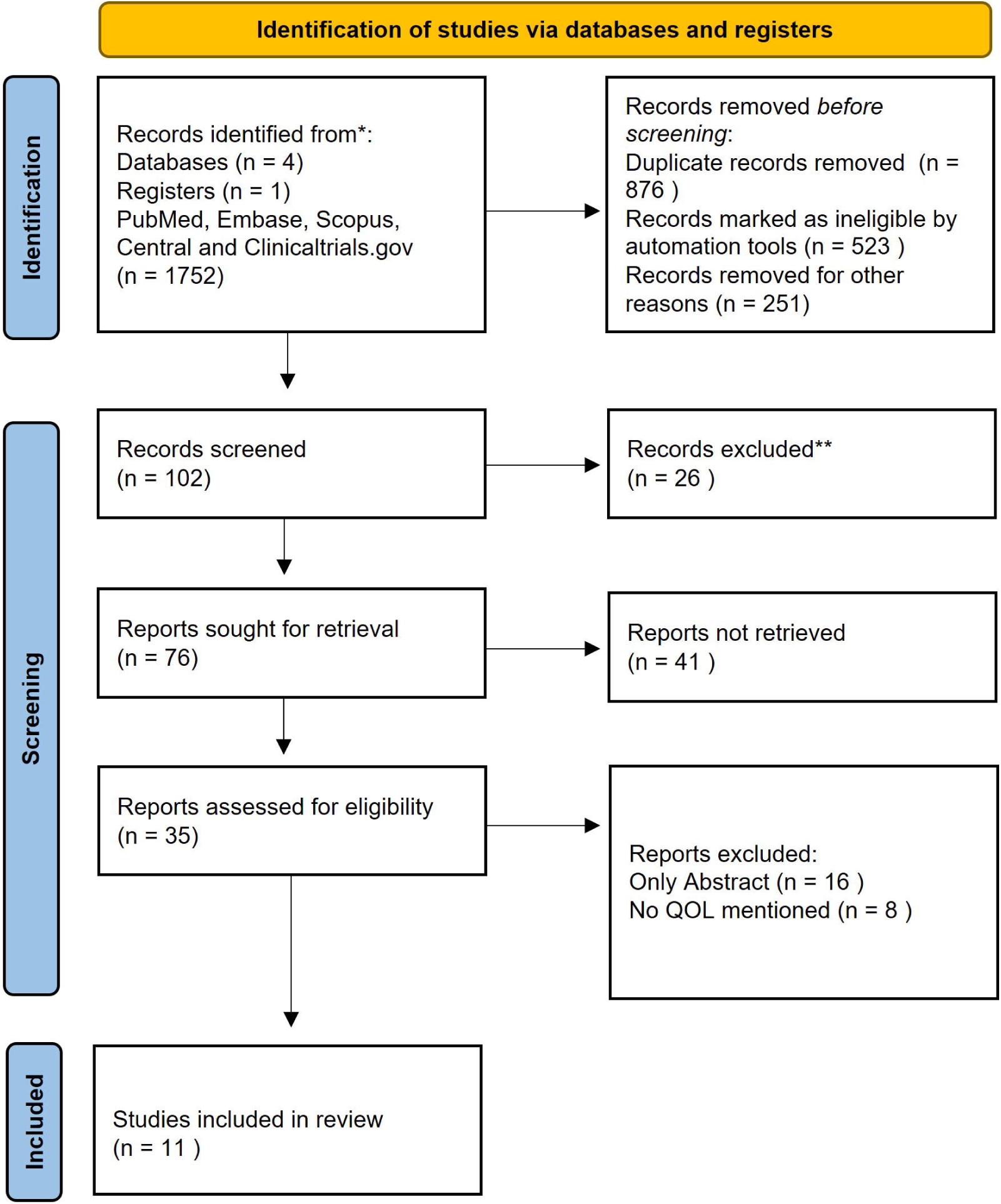
PRISMA Flow Diagram

**Figure 2.**
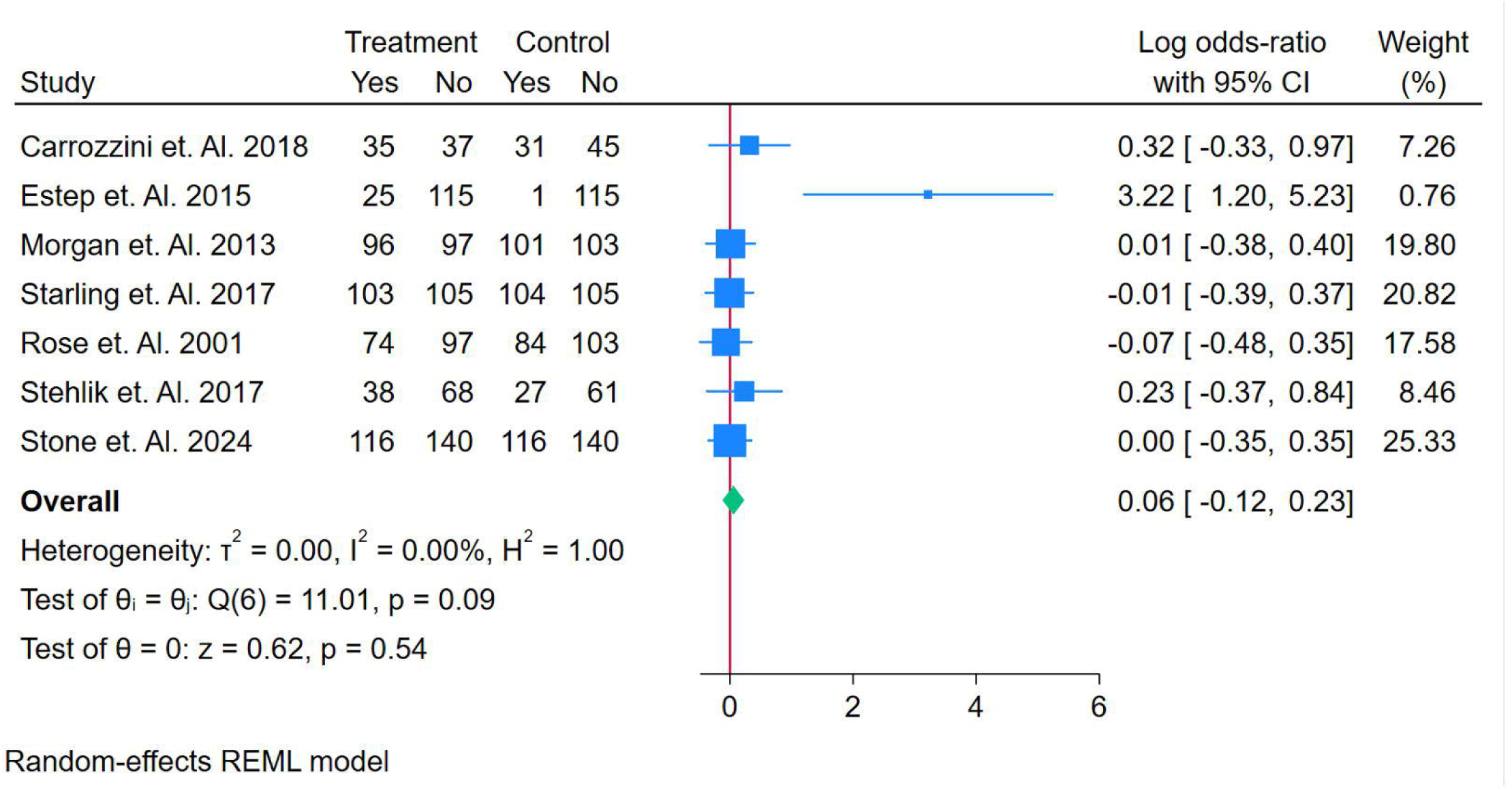
Forest Plot of Odds-ratio of Survival Outcomes after 1 Month of LVAD Implantation

**Figure 3.**
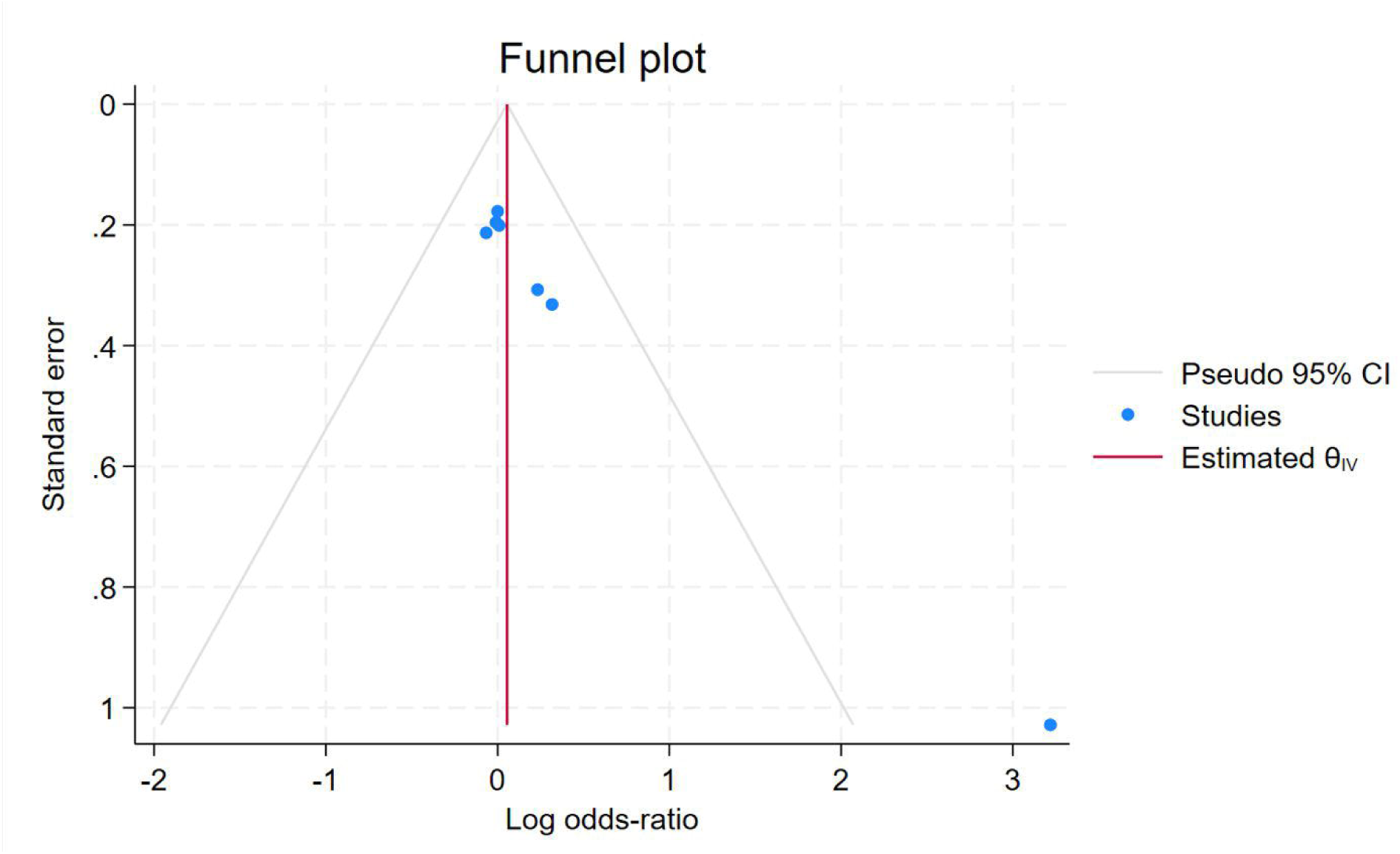
Funnel Plot of Odds-ratio of Survival Outcomes after 1 Month of LVAD Implantation

#### Survival in Two-month post LVAD

Survival outcomes after 2 months of Left Ventricular Assist Device (LVAD) implantation, based on data from four studies. The results from these studies vary, with some suggesting a modest effect and others showing no significant impact. **Carrozzini et al. (2018)** [14] reported an odds-ratio of 0.38, with a confidence interval of [− 0.27, 1.03], contributing 14.68% to the overall weight. While this suggests a possible positive effect, the confidence interval includes 1, indicating no statistical significance. **Morgan et al. (2013)**, [17] contributing the largest weight (39.97%), found an odds-ratio of 0.01 with a confidence interval of [−0.39, 0.40], indicating no meaningful effect on survival outcomes. **Rose et al. (2001)** [19] reported an odds-ratio of −0.07 with a 95% confidence interval of [−0.48, 0.35], contributing 35.67% to the overall estimate, which also suggests no significant effect. Lastly, **Stehlik et al. (2017)** [20] showed an odds-ratio of 0.58 with a confidence interval of [− 0.22, 1.39], contributing 9.68% to the pooled estimate, but the confidence interval includes 1, indicating no clear effect. Figure 4,5 and Table S3.

**Figure 4.**
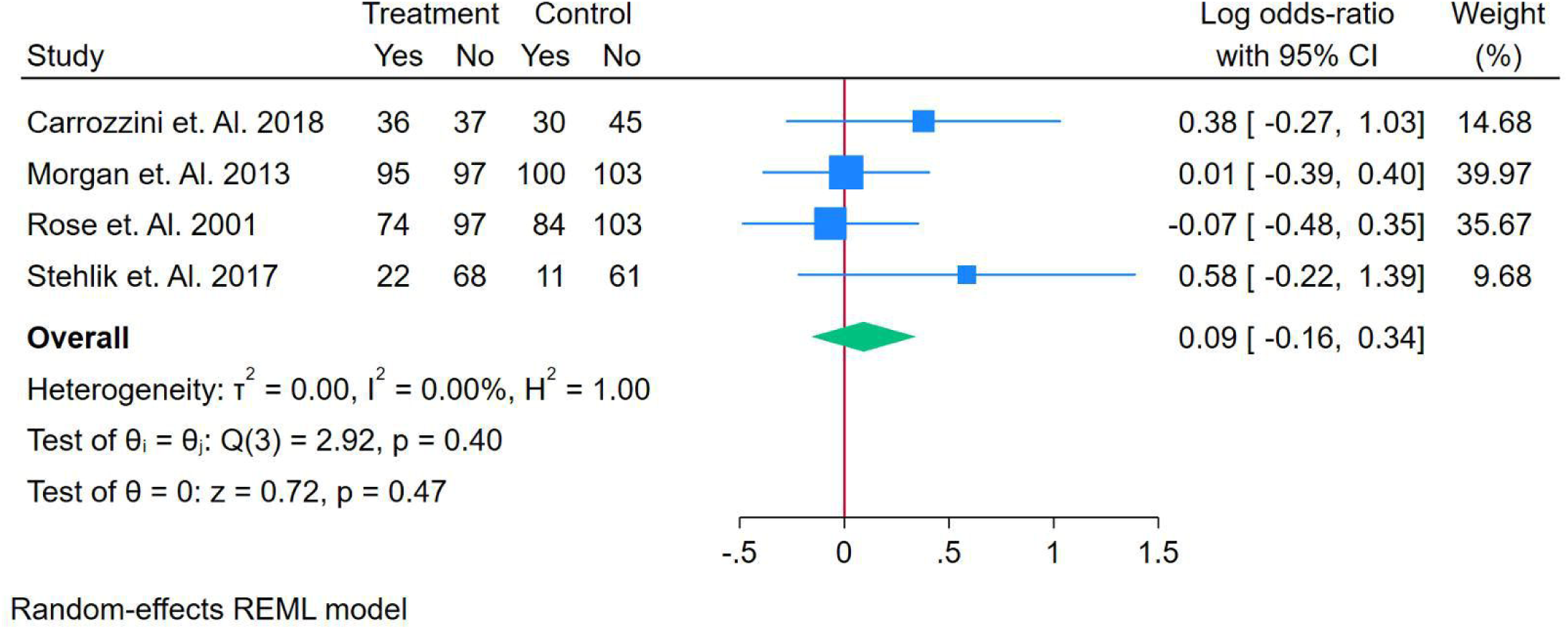
Forest Plot of Odds-ratio of Survival Outcomes after 2 Month of LVAD Implantation

**Figure 5.**
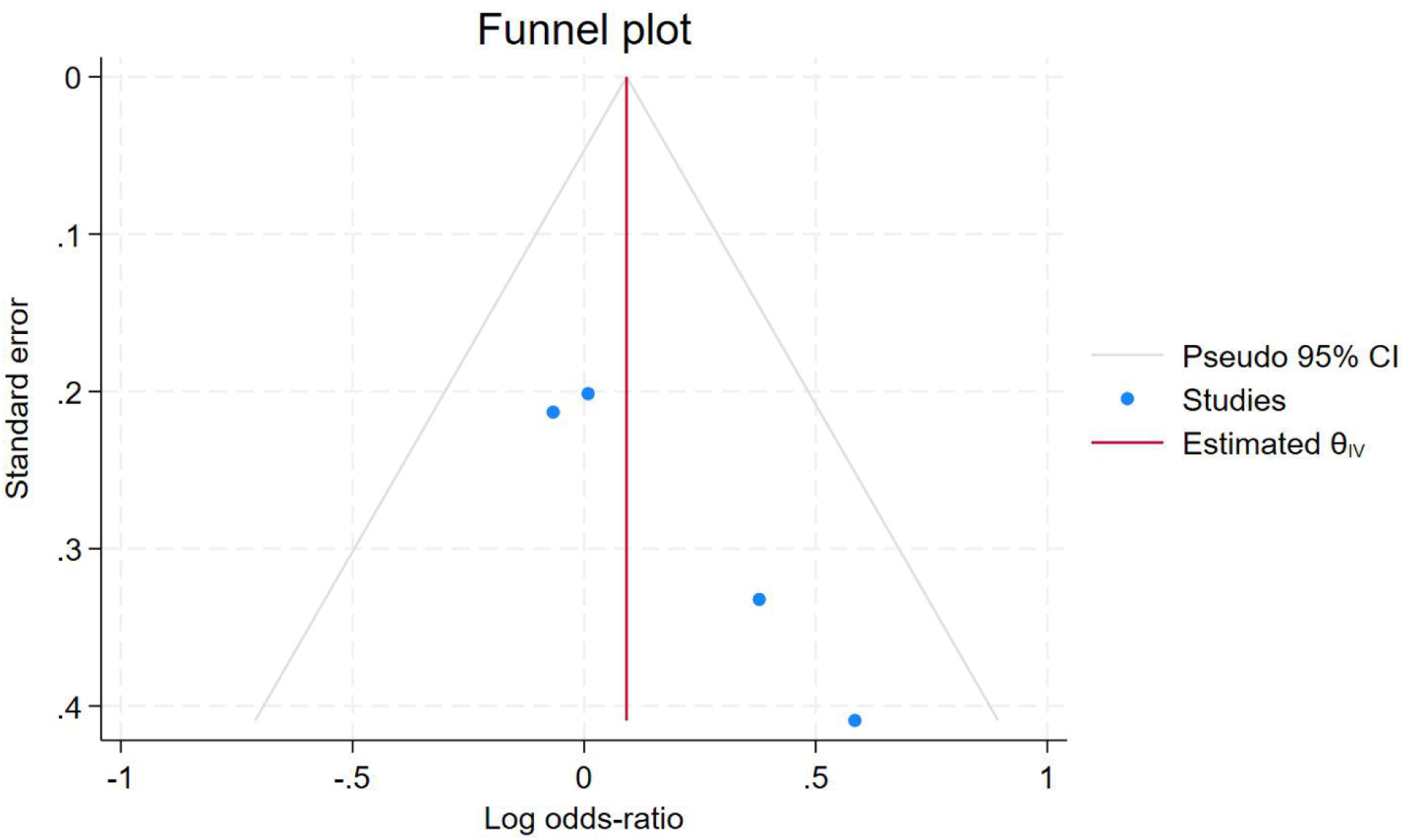
Funnel Plot of Odds-ratio of Survival Outcomes after 1 Month of LVAD Implantation

The overall pooled odds-ratio was 0.09, with a 95% confidence interval of [−0.16, 0.34], which suggests no statistically significant effect of LVAD implantation on survival outcomes after 2 months, as the confidence interval includes 0. The **heterogeneity statistics** (τ² = 0.00, I² = 0.00%, H² = 1.00) show no variability between the studies, indicating that the results are highly consistent. The **homogeneity test** (Q(3) = 2.92, p = 0.40) further confirms that there are no significant differences between the studies, and the **test for overall effect** (z = 0.72, p = 0.47) shows no significant pooled effect. These findings collectively suggest that LVAD implantation does not have a clear or significant impact on survival outcomes at 2 months, and the consistency across studies strengthens this conclusion.

#### Survival in Six-month post LVAD

Survival outcomes after 6 months of Left Ventricular Assist Device (LVAD) implantation, based on data from five studies. The odds-ratios from the studies vary, with some suggesting a potential effect while others show no impact. **Carrozzini et al. (2018)** [14] reported an odds-ratio of 0.46 with a confidence interval of [−0.21, 1.12], contributing 10.25% to the overall weight. While this suggests a possible positive effect, the confidence interval includes 1, indicating no statistical significance. **Morgan et al. (2013) [17]** found an odds-ratio of 0.10 with a confidence interval of [−0.30, 0.51], contributing 27.89% to the pooled estimate, and this result suggests no significant effect on survival outcomes. **Starling et al. (2017) [18]** showed an odds-ratio of 0.00 with a confidence interval of [−0.39, 0.39], contributing 29.52%, indicating no significant effect. **Rose et al. (2001) [19]** reported an odds-ratio of 0.02 with a confidence interval of [−0.40, 0.44], contributing 25.31%, and similarly found no significant impact. Lastly, **Stehlik et al. (2017)** [20] reported an odds-ratio of 0.58 with a confidence interval of [−0.22, 1.39], contributing 7.03%, showing a possible positive effect, but the confidence interval includes 1, indicating no clear benefit. Figure 6,7 and Table S4.

**Figure 6.**
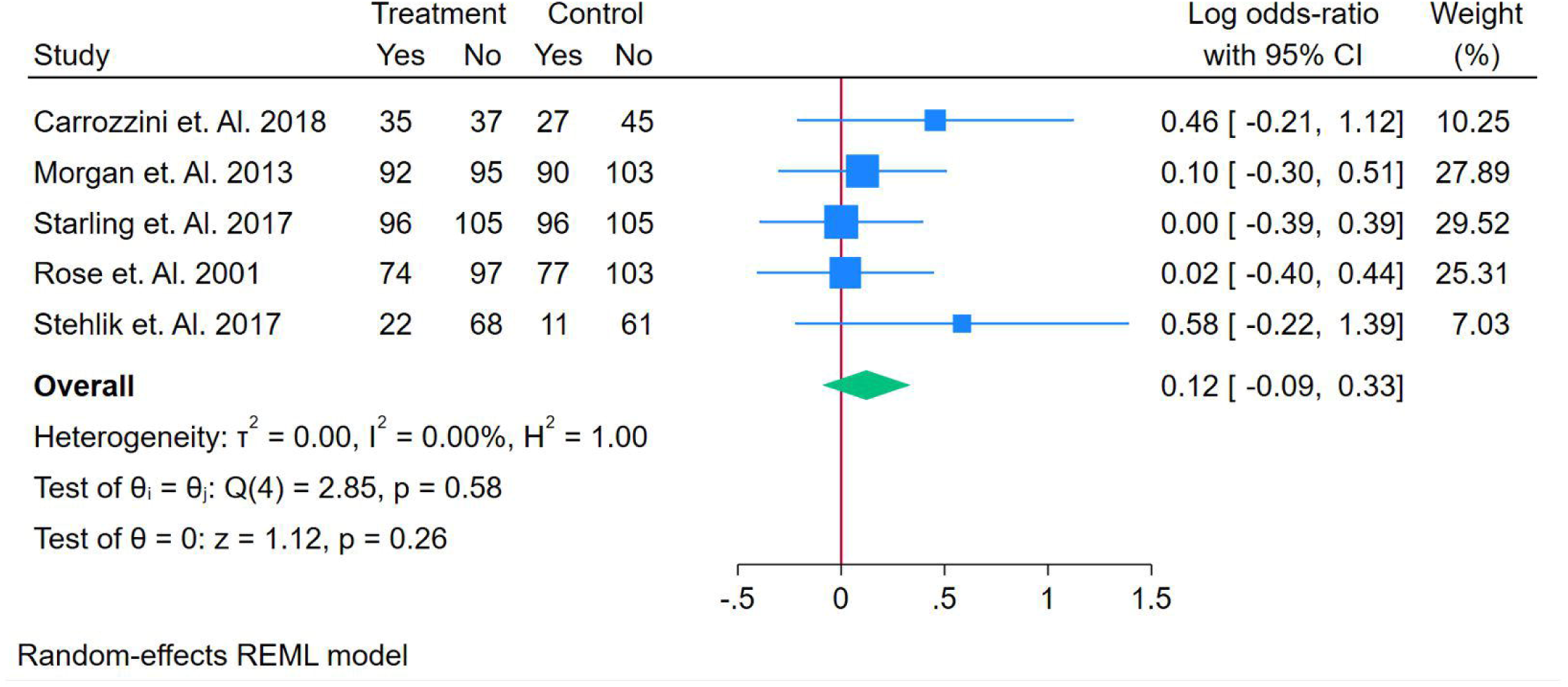
Forest Plot of Odds-ratio of Survival Outcomes after 6 Month of LVAD Implantation

**Figure 7.**
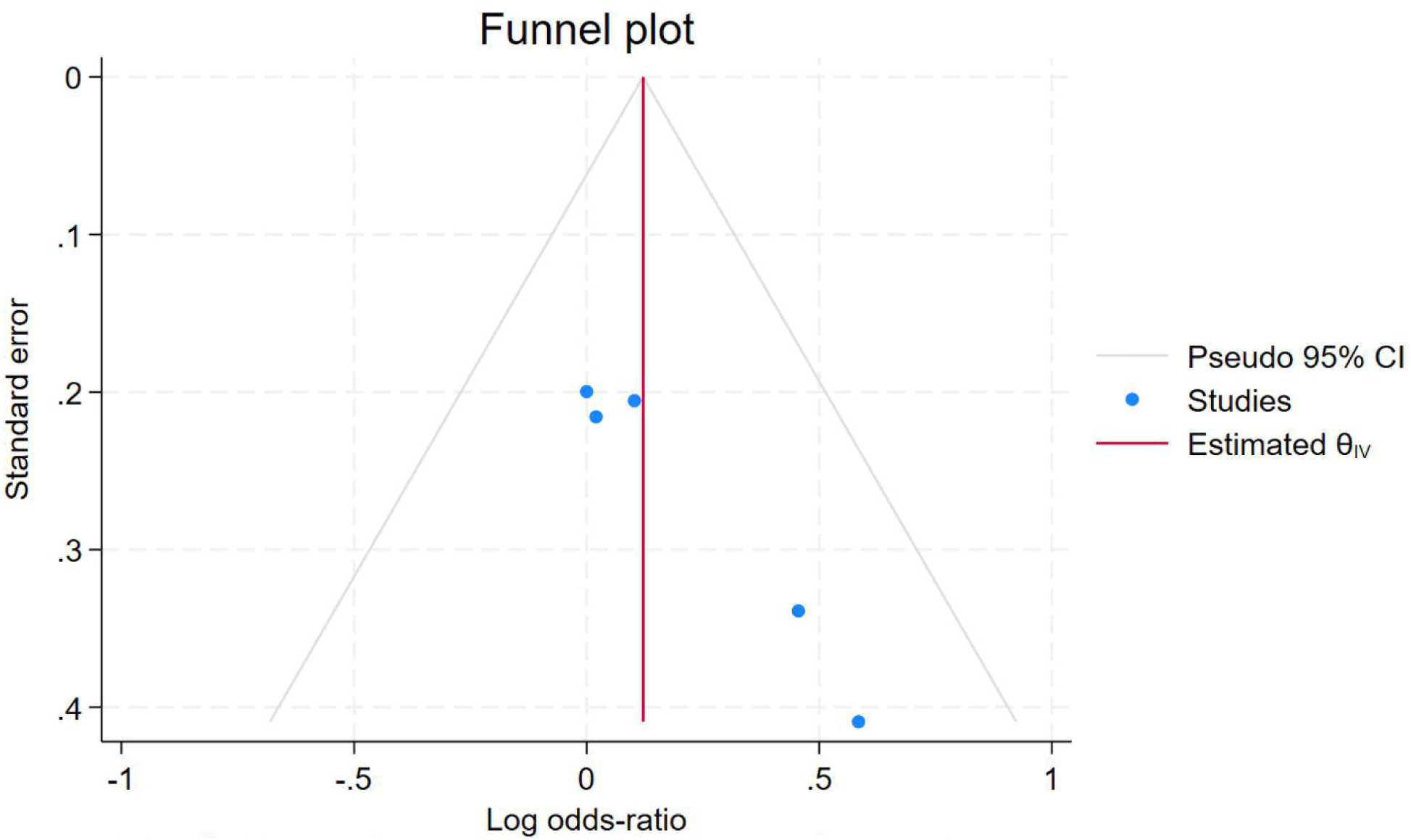
Funnel Plot of Odds-ratio of Survival Outcomes after 6 Month of LVAD Implantation

The overall pooled odds-ratio is 0.12, with a 95% confidence interval of [−0.09, 0.33], suggesting no statistically significant effect of LVAD implantation on survival outcomes after 6 months, as the confidence interval includes 0. The **heterogeneity analysis** (τ² = 0.00, I² = 0.00%, H² = 1.00) shows no heterogeneity across the studies, indicating consistent results. The **homogeneity test** (Q(4) = 2.85, p = 0.58) further supports this, showing no significant variability between the studies. Finally, the **test for overall effect** (z = 1.12, p = 0.26) confirms that the pooled estimate does not show a significant effect. Overall, these findings suggest that LVAD implantation does not significantly improve survival outcomes at 6 months, and the consistency across studies strengthens this conclusion.

#### Survival in Twelve-month post LVAD

Survival outcomes after 12 months of Left Ventricular Assist Device (LVAD) implantation, based on data from eight studies. The results show some variability across the studies, with several suggesting no significant effect, while others indicate potential benefits. **Carrozzini et al. (2018)** [14] reported an odds-ratio of 0.54 with a confidence interval of [−0.14, 1.22], contributing 5.11% to the overall weight. Although this suggests a potential positive effect, the confidence interval includes 1, indicating no statistical significance. **Hasin et al. (2013)** [15] found an odds-ratio of 0.06 with a confidence interval of [−0.46, 0.58], contributing 8.75%, showing no significant effect on survival. **Estep et al. (2015)** [16] reported an odds-ratio of 0.51 with a confidence interval of [0.06, 0.96], contributing 11.61%, indicating a possible positive effect with the confidence interval excluding 0. **Morgan et al. (2013)** [17] found an odds-ratio of 0.30 with a confidence interval of [−0.14, 0.75], contributing 12.11%, suggesting a moderate positive effect, although it is not statistically significant as the confidence interval includes 0. **Starling et al. (2017)** [18] and **Rose et al. (2001) [19]** reported odds-ratios of 0.00 and 0.09, respectively, with confidence intervals overlapping 0, indicating no effect. **Stehlik et al. (2017) [20]** reported an odds-ratio of 0.40 with a wide confidence interval of [−1.07, 1.87], contributing 1.09%, suggesting a possible positive effect, but the wide range includes no effect. Lastly, **Takeda et al. (2014)** [21] showed an odds-ratio of 0.02 with a confidence interval of [−0.39, 0.44], contributing 13.79%, indicating no significant effect on survival. Figure 8,9 and Table S5.

**Figure 8.**
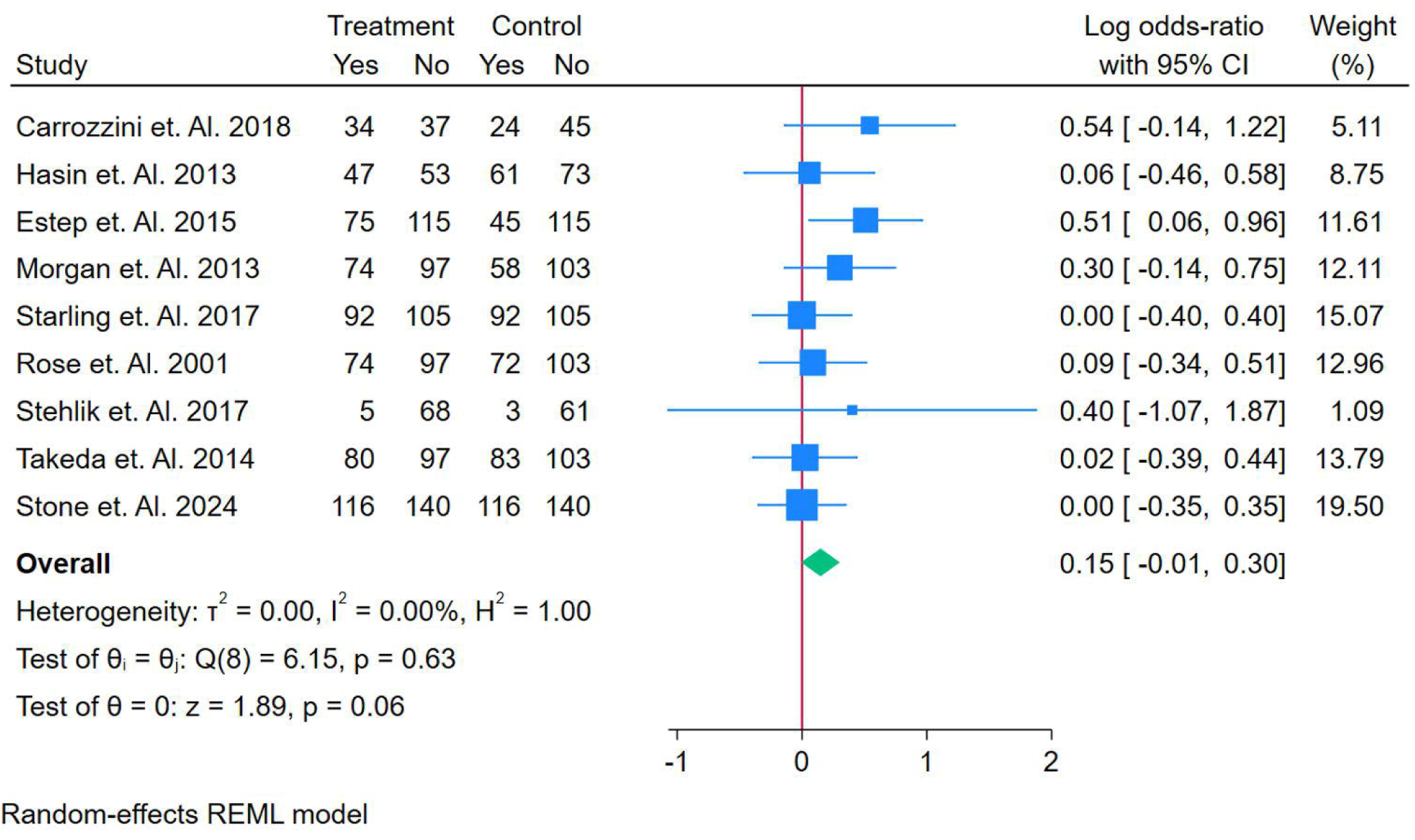
Forest Plot of Odds-ratio of Survival Outcomes after 12 Month of LVAD Implantation

**Figure 9.**
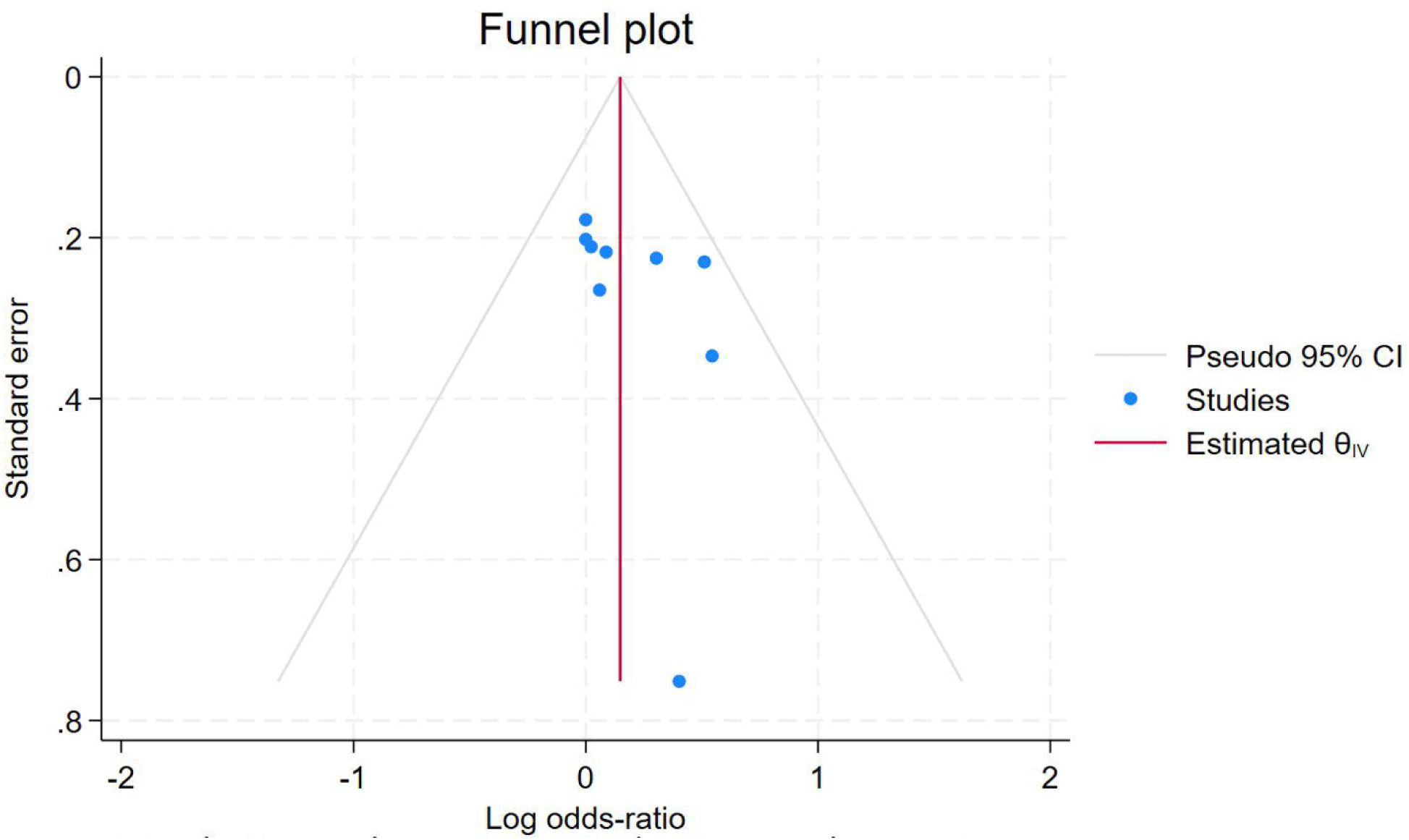
Funnel Plot of Odds-ratio of Survival Outcomes after 12 Month of LVAD Implantation

The overall pooled odds-ratio is 0.15, with a 95% confidence interval of [−0.01, 0.30], suggesting no statistically significant effect of LVAD implantation on survival outcomes at 12 months, as the confidence interval includes 0. The **heterogeneity statistics** (τ² = 0.00, I² = 0.00%, H² = 1.00) indicate no variation across the studies, showing consistent results. The **homogeneity test** (Q(8) = 6.15, p = 0.63) further confirms no significant heterogeneity, and the **test for overall effect** (z = 1.89, p = 0.06) shows a borderline effect, which does not reach statistical significance (p < 0.05). In conclusion, although there is some variation across individual studies, the overall pooled estimate suggests that LVAD implantation does not significantly affect survival outcomes at 12 months, and the consistency across studies strengthens this finding.

## Adverse Events

### Death events in LVADs

Risk ratio of death as an adverse effect following Left Ventricular Assist Device (LVAD) implantation, based on data from ten studies. The individual study results show some variability, with a few studies suggesting potential protective effects, while others show no significant impact on mortality. **Ammirati et al. (2016) [13]** reported a risk-ratio of −0.46, indicating a possible protective effect, but the confidence interval overlaps zero, suggesting no statistical significance. **Carrozzini et al. (2018)** [14] also found a risk-ratio of −0.33, again indicating a non-significant protective effect. **Hasin et al. (2013)** [15] reported a risk-ratio of 0.20, suggesting a potential benefit, although the confidence interval includes zero, pointing to an uncertain conclusion. **Estep et al. (2015)** [16] showed a risk-ratio of 0.00, indicating no effect on mortality. **Morgan et al. (2013)** [17] and **Starling et al. (2017)** [18] found risk-ratios of −0.19 and −0.22, respectively, suggesting no significant impact on mortality. **Rose et al. (2001)** showed a similar result with a risk-ratio of −0.22, while **Stehlik et al. (2017) [20]** and **Takeda et al. (2014)** [21] reported risk-ratios of −0.09 and 0.00, respectively, again showing no significant effect. **Stone et al. (2024)** found a risk-ratio of 0.26, suggesting a potential positive effect, but the wide confidence interval includes no effect. Figure 10, 11 and Table S6.

**Figure 10.**
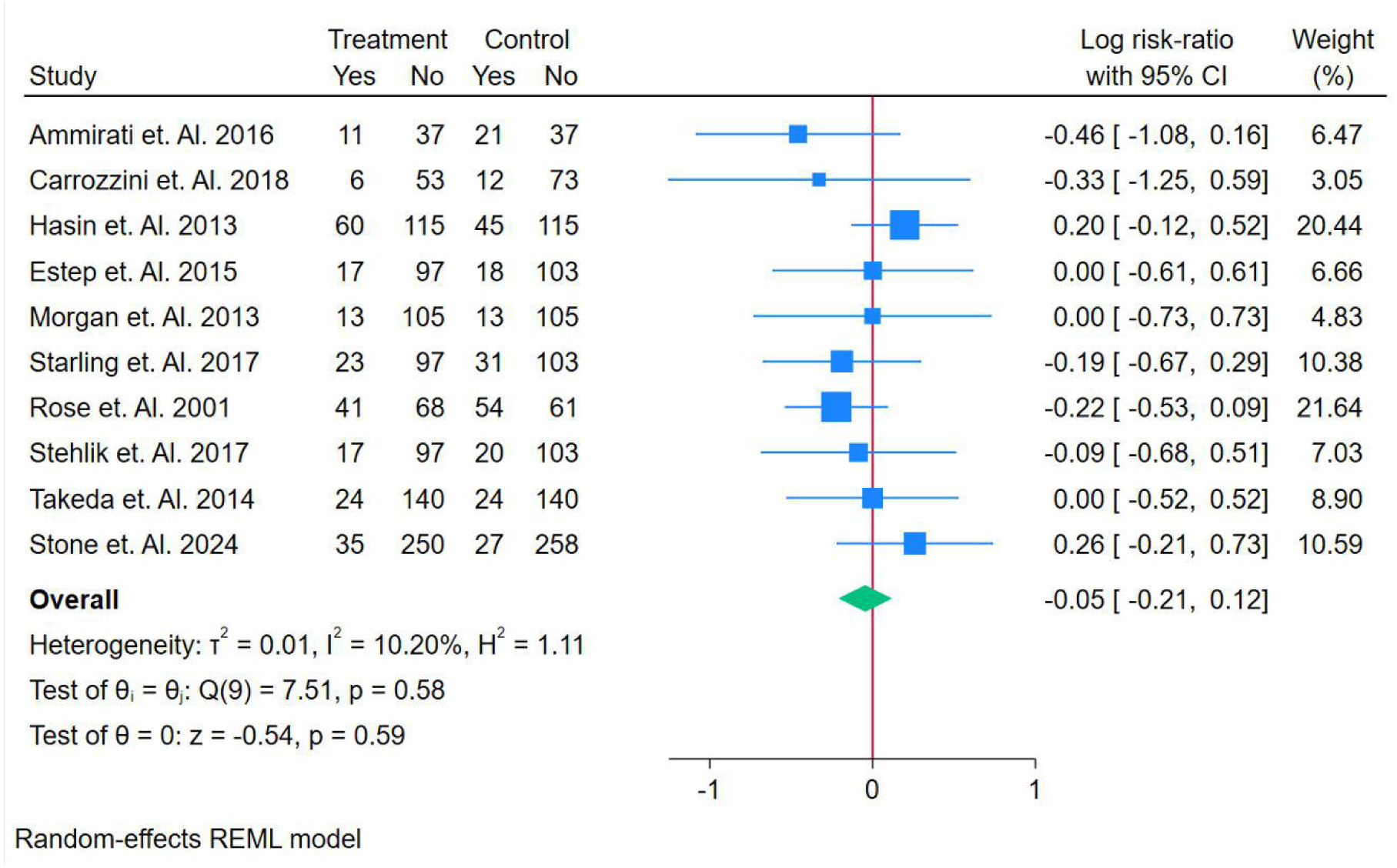
Forest Plot of Risk Ratio of Death as Adverse Effects post LVAD Implantation

**Figure 11.**
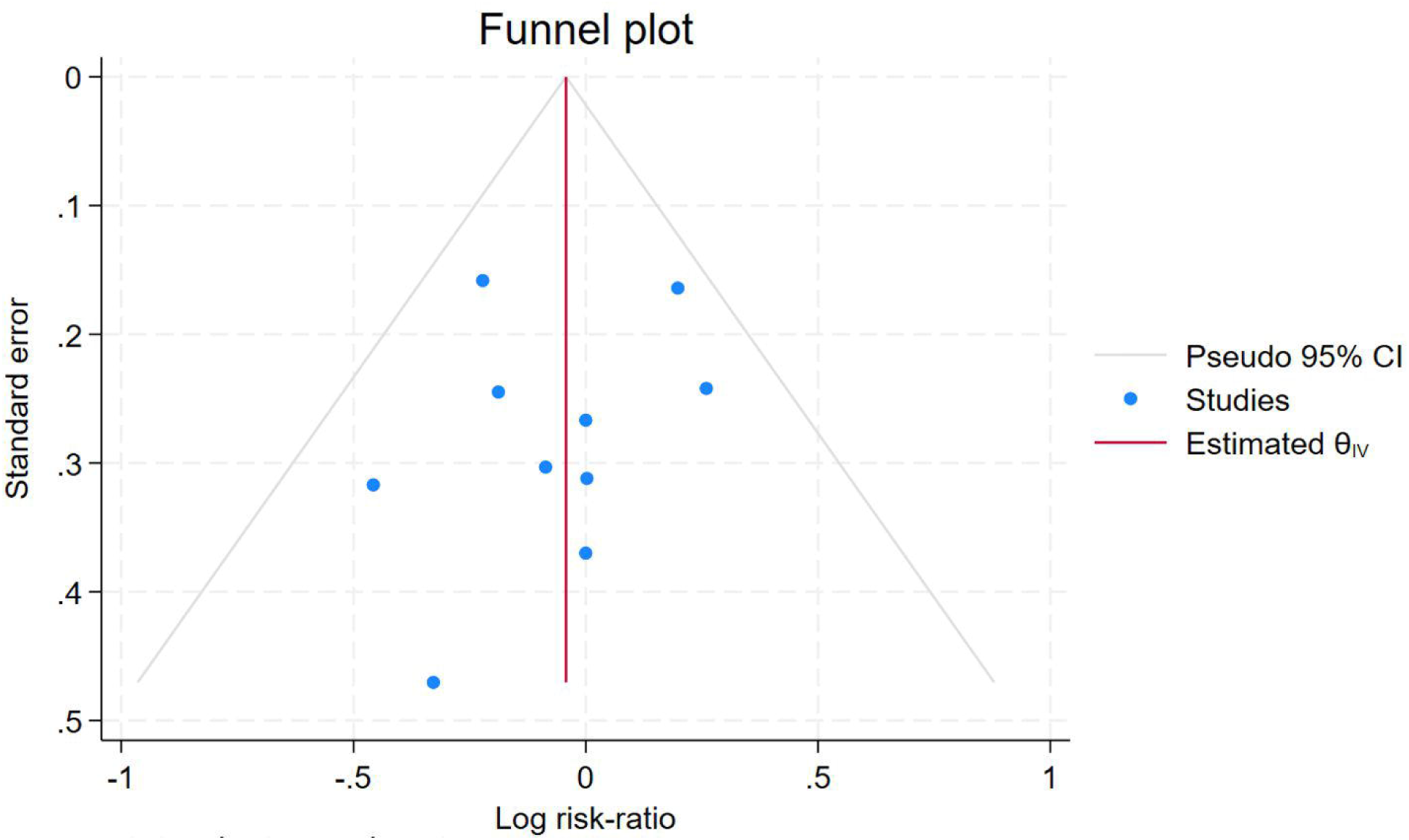
Funnel Plot of Risk ratio of Death post LVAD Implant

The overall pooled risk-ratio is −0.05, with a 95% confidence interval of [−0.21, 0.12], indicating no significant reduction in the risk of death following LVAD implantation, as the confidence interval includes zero. The **heterogeneity statistics** (τ² = 0.01, I² = 10.20%, H² = 1.11) suggest low variability between the studies, indicating consistent results. The **homogeneity test** (Q(9) = 7.51, p = 0.58) confirms no significant heterogeneity, suggesting that the differences observed across studies are due to random error. Finally, the **test for overall effect** (z = −0.54, p = 0.59) shows no statistical significance, further supporting the conclusion that LVAD implantation does not significantly reduce the risk of death. Overall, while some individual studies show potential benefits, the pooled estimate and the consistency across studies suggest no clear effect on mortality.

### Thrombotic Events

thrombotic events as adverse effects following Left Ventricular Assist Device (LVAD) implantation, based on data from ten studies. The individual studies show some variability in their findings, with a few suggesting potential protective effects, while others indicate no significant impact. **Ammirati et al. (2016)** [13] reported a risk-ratio of −0.46 with a confidence interval of [−1.08, 0.16], suggesting a possible protective effect against thrombotic events, though the confidence interval includes zero, indicating no statistical significance. **Carrozzini et al. (2018) [14]** found a similar result with a risk-ratio of −0.33, and **Hasin et al. (2013) [15]** reported a risk-ratio of 0.20, both indicating non-significant effects as their confidence intervals overlap zero. **Estep et al. (2015) [16]** and **Morgan et al. (2013)** [17] found a risk-ratio of 0.00, suggesting no effect on thrombotic events, with confidence intervals spanning from negative to positive values. **Starling et al. (2017)**, **Rose et al. (2001)**, and **Stehlik et al. (2017)** [18–20] also reported risk-ratios close to zero, indicating no significant effect. **Takeda et al. (2014)** found a risk-ratio of 0.00, suggesting no impact, and **Stone et al. (2024) [22]** reported a risk-ratio of 0.26, indicating a possible positive effect, but the confidence interval includes zero, suggesting uncertainty. Figure 12, 13, and Table S7.

**Figure 12.**
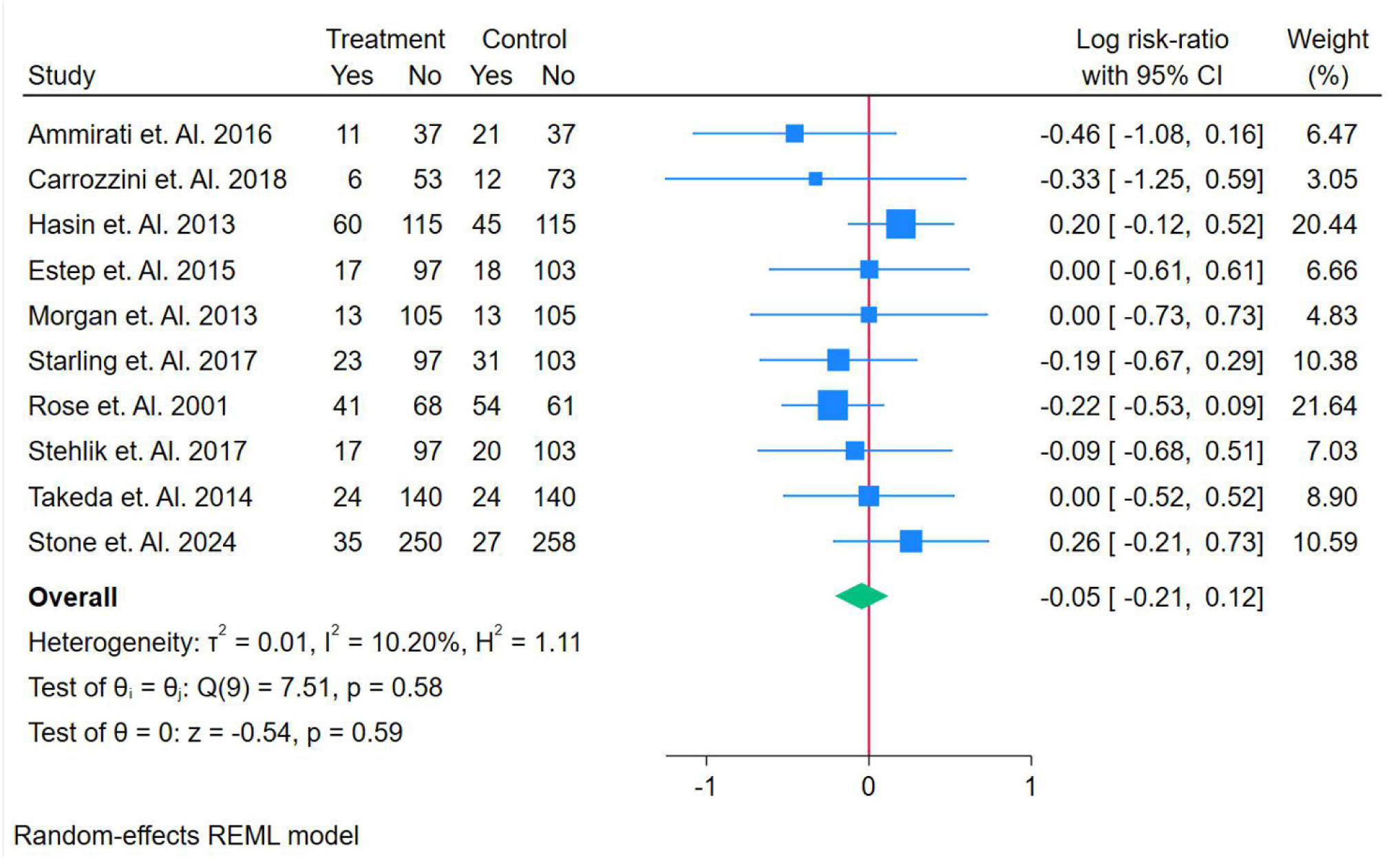
Forest Plot of Risk Ratio of Thrombotic as Adverse Effects post LVAD Implantation

**Figure 13.**
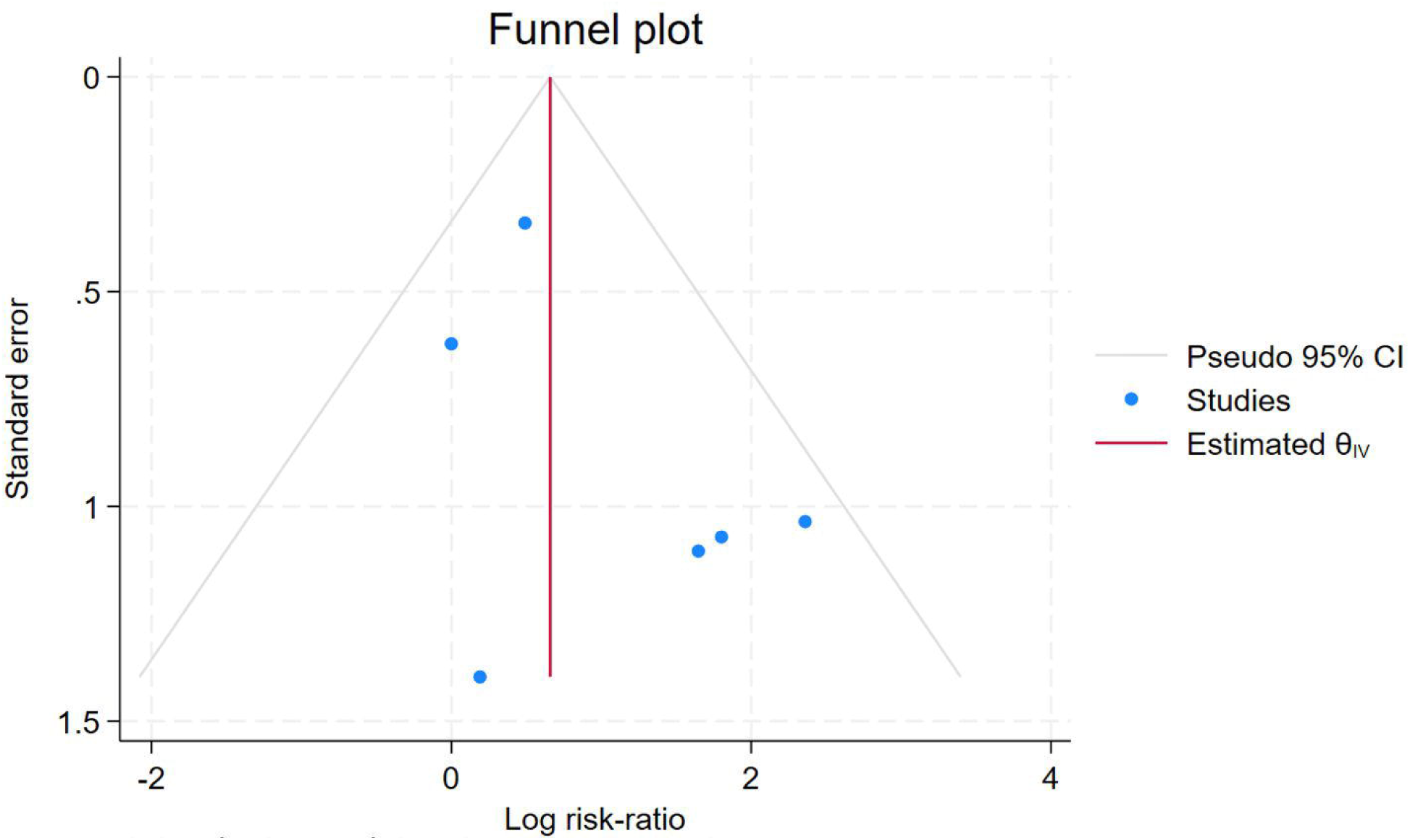
Funnel Plot of Risk ratio of Thrombotic post LVAD Implant

The overall pooled risk-ratio is −0.05, with a 95% confidence interval of [−0.21, 0.12], indicating no significant effect of LVAD implantation on thrombotic events, as the confidence interval includes zero. The **heterogeneity statistics** (τ² = 0.01, I² = 10.20%, H² = 1.11) indicate low variability across the studies, suggesting consistent results. The **homogeneity test** (Q(9) = 7.51, p = 0.58) confirms no significant heterogeneity, indicating the differences observed across studies are likely due to random error. The **test for overall effect** (z = −0.54, p = 0.59) further supports that there is no statistically significant effect of LVAD implantation on thrombotic events. Overall, despite some variation in individual study results, the pooled estimate suggests that LVAD implantation does not significantly affect the occurrence of thrombotic events, with the confidence interval including zero and minimal heterogeneity across studies.

### Bleeding Events

Bleeding as an adverse effect following Left Ventricular Assist Device (LVAD) implantation, based on data from seven studies. The individual studies show some variation in their results, with several indicating a significant increase in the risk of bleeding. **Ammirati et al. (2016)** [13] reported a risk-ratio of 1.24 with a confidence interval of [−0.98, 3.46], suggesting a possible increase in bleeding risk, but the confidence interval includes 1, indicating no statistical significance. **Carrozzini et al. (2018)** [14] found a significant risk-ratio of 2.61 with a confidence interval of [0.60, 4.63], indicating a clear increase in bleeding risk. Similarly, **Hasin et al. (2013)** [15] reported a risk-ratio of 2.66 with a confidence interval of [1.53, 3.80], indicating a significant increase in bleeding risk, and **Estep et al. (2015)** [16] reported a risk-ratio of 3.48 with a confidence interval of [1.51, 5.45], showing a significant increase. **Starling et al. (2017)** [18] and **Rose et al. (2001) [19]** also found increased bleeding risks, with risk-ratios of 2.01 and 2.03, respectively, though Rose’s confidence interval includes 1, suggesting a less certain effect. **Takeda et al. (2014)** [21] found no effect with a risk-ratio of 0.00 and a confidence interval of [−0.55, 0.55], suggesting no significant change in bleeding risk. Figure 14, 15 and Table S8.

**Figure 14.**
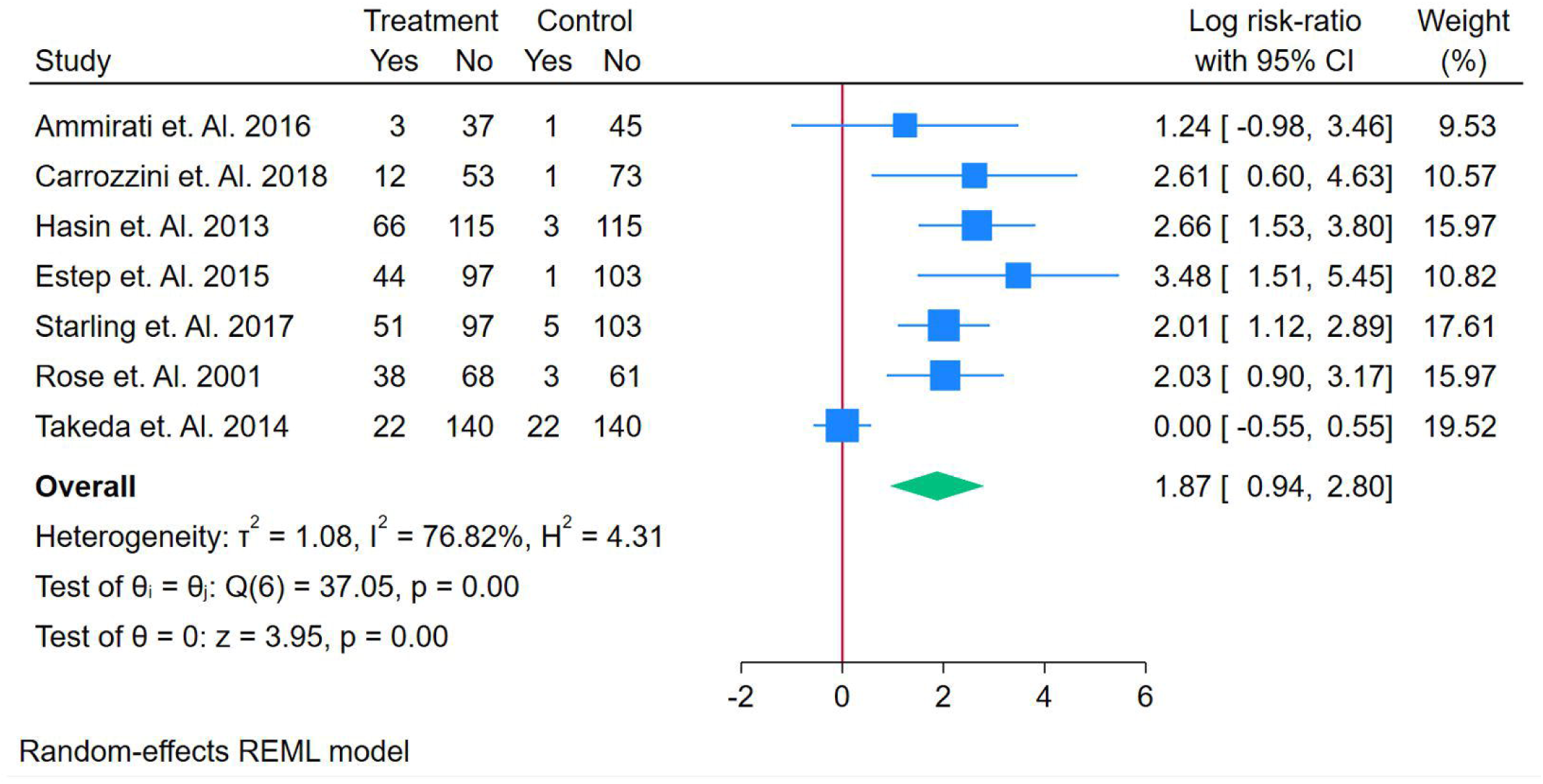
Forest Plot of Risk Ratio of Bleeding as Adverse Effects post LVAD Implantation

**Figure 15.**
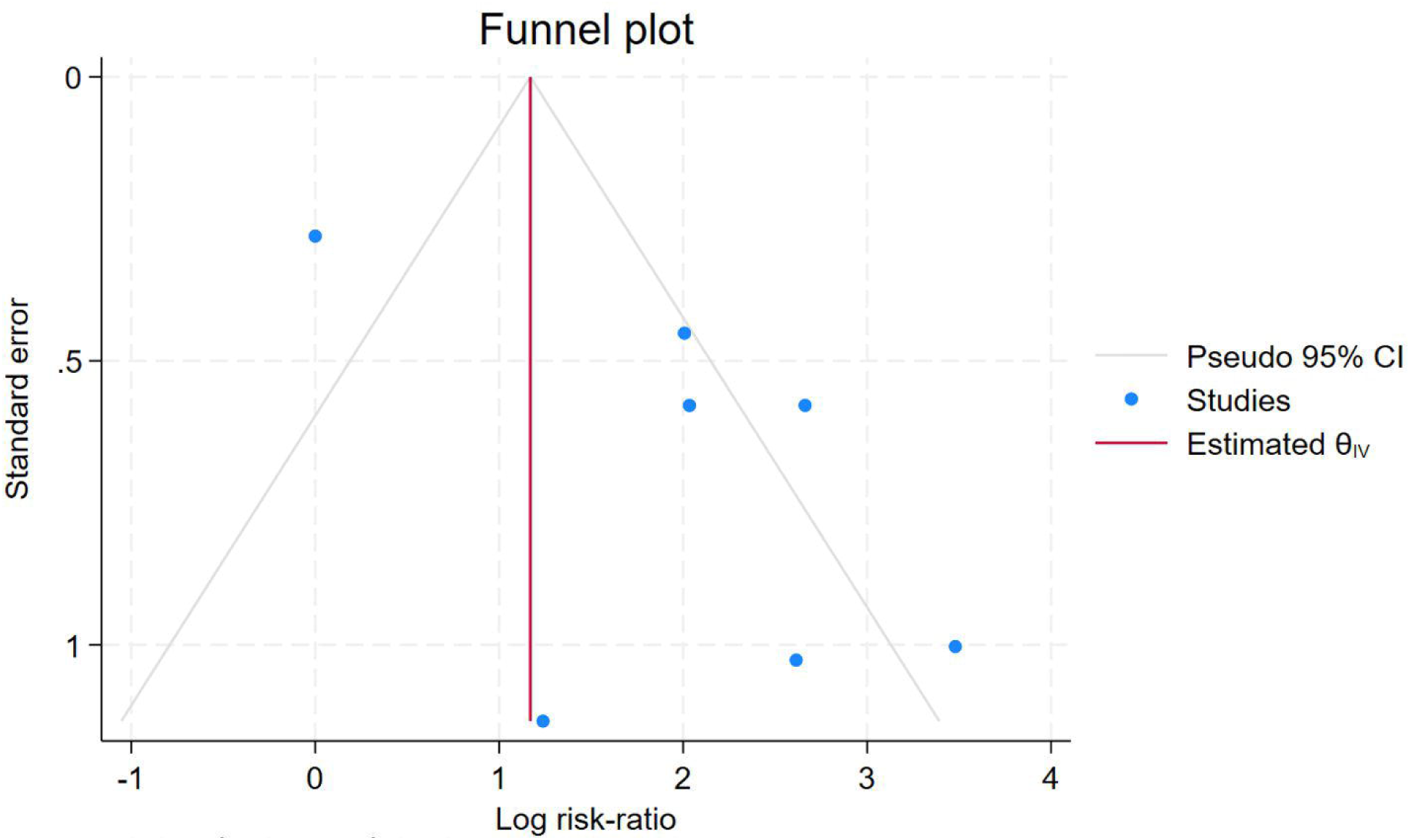
Funnel Plot of Risk ratio of Bleeding post LVAD Implant

The overall pooled risk-ratio is 1.87, with a 95% confidence interval of [0.94, 2.80], indicating a significant increase in bleeding risk post-LVAD implantation. The **heterogeneity statistics** (τ² = 1.08, I² = 76.82%, H² = 4.31) indicate substantial variation across the studies, suggesting that factors other than the LVAD implantation itself may influence the outcomes. The **homogeneity test** (Q(6) = 37.05, p = 0.00) confirms the presence of significant heterogeneity, meaning that the studies are not entirely consistent. Despite this variability, the **test for overall effect** (z = 3.95, p = 0.00) shows a statistically significant result, confirming that LVAD implantation is associated with an increased risk of bleeding. Overall, while there is variation across the studies, the pooled estimate suggests that LVAD implantation significantly increases the risk of bleeding, although the high heterogeneity points to other contributing factors in these outcomes.

### Neurological Consequences

Neurological consequences as adverse effects following Left Ventricular Assist Device (LVAD) implantation, based on data from six studies. The individual studies show variability in their findings. **Ammirati et al. (2016)** reported a risk-ratio of 0.18 with a confidence interval of [−1.36, 1.73], suggesting a very small potential effect, though the confidence interval includes zero, indicating no statistical significance. **Carrozzini et al. (2018)** [14] found a risk-ratio of 1.66 with a confidence interval of [−0.49, 3.82], suggesting a possible positive effect, but the confidence interval includes 1, meaning the result is not definitive. **Estep et al. (2015)** [16] reported a risk-ratio of 1.39 with a confidence interval of [−0.14, 2.91], indicating a potential positive effect, but again, the confidence interval includes 1. **Starling et al. (2017)** [18] showed a risk-ratio of 1.00 with a confidence interval of [−0.11, 2.12], indicating no significant effect. **Rose et al. (2001) [19]** reported a risk-ratio of 1.30 with a confidence interval of [0.39, 2.20], suggesting a potential increase in neurological consequences. **Takeda et al. (2014)** [21] found a risk-ratio of 0.00 with a confidence interval of [−0.90, 0.90], indicating no effect. Figure 16, 17 and Table S9.

**Figure 16.**
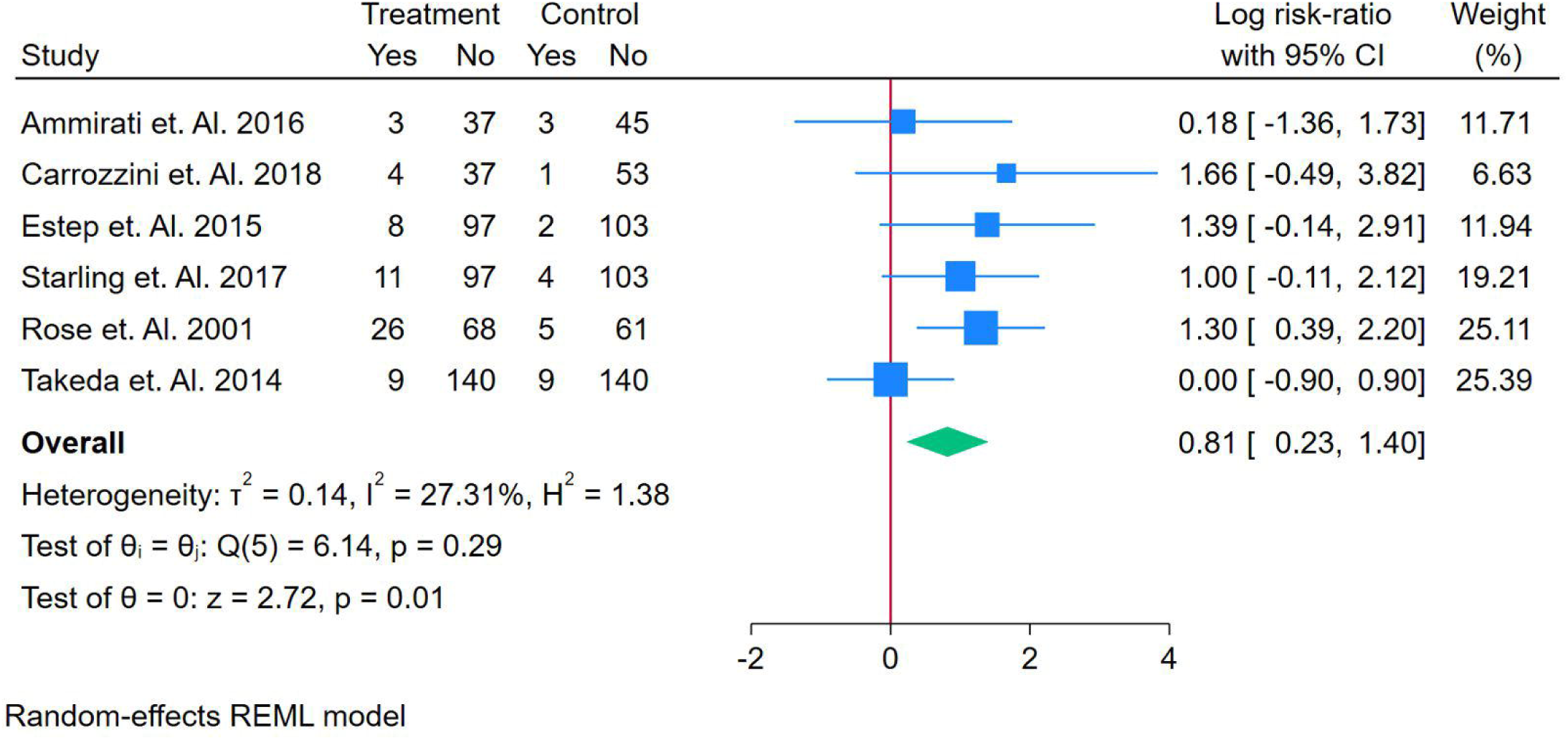
Forest Plot of Risk Ratio of Neurological Consequences as Adverse Effects post LVAD Implantation

**Figure 17.**
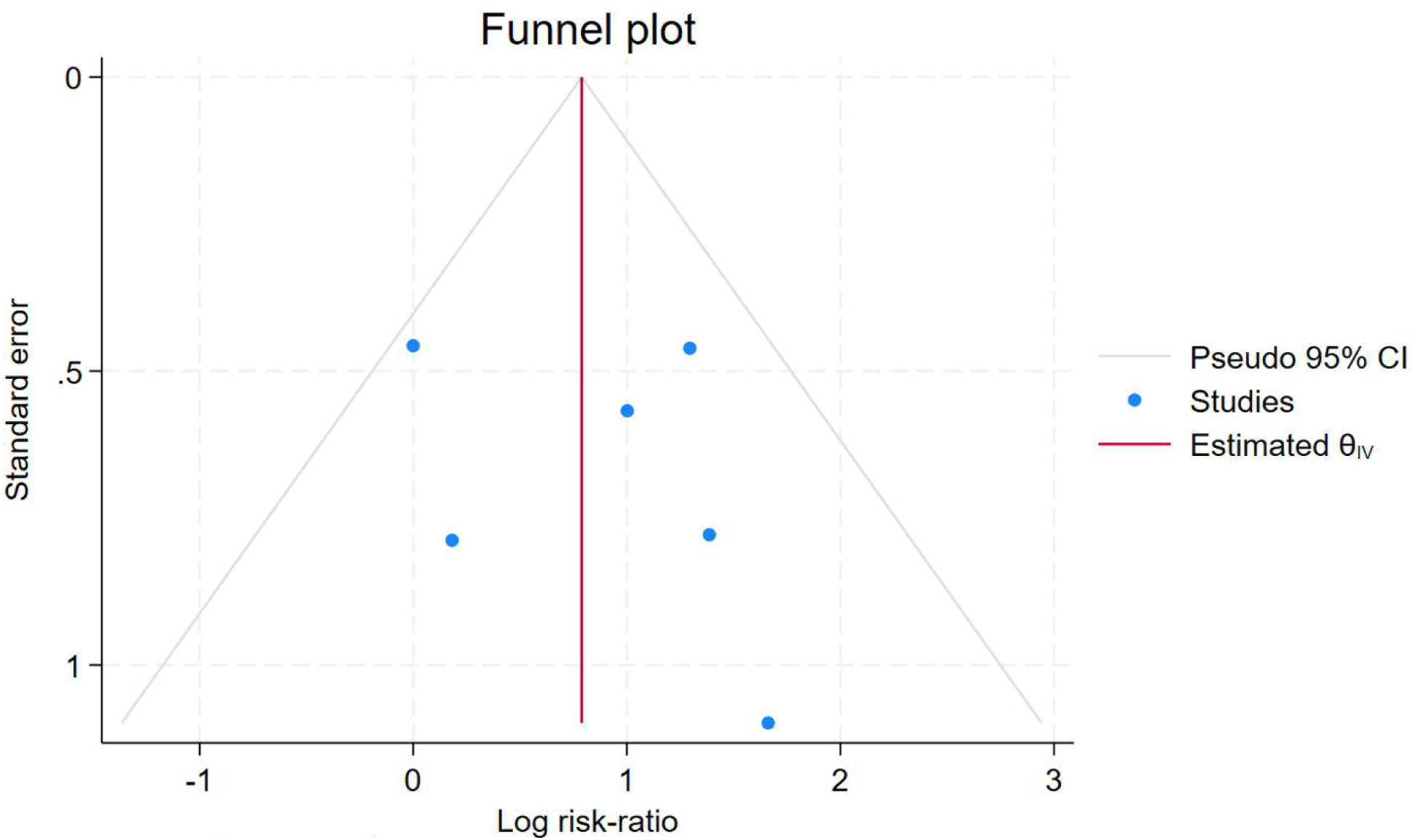
Funnel Plot of Risk ratio of Neurological consequences post LVAD Implant

The overall pooled risk-ratio is 0.81, with a 95% confidence interval of [0.23, 1.40], suggesting a possible increase in neurological consequences after LVAD implantation. However, the confidence interval includes 1, indicating that this result is not statistically significant. The **heterogeneity statistics** (τ² = 0.14, I² = 27.31%, H² = 1.38) suggest some variability across the studies, but overall, the results are relatively consistent. The **homogeneity test** (Q(5) = 6.14, p = 0.29) indicates no significant heterogeneity, meaning the variation across studies is likely due to random error. The **test for overall effect** (z = 2.72, p = 0.01) shows a statistically significant result, suggesting a slight increase in neurological consequences.

### Quality of Life

The data highlights improvements in quality of life (QOL) for patients receiving Left Ventricular Assist Device (LVAD) treatment, as assessed through various health-related measures. The EQ-5D scores reported by Starling et al. (2017) and Stehlik et al. (2017) [20] show moderate to significant improvements in general health-related QOL, with scores of 27 ± 24 and 50 ± 20, respectively. This indicates enhanced overall health status following LVAD implantation. The PHQ-9 scores revealed moderate depressive symptoms, with Starling et al. (2017) [18] reporting an average of 4.6 ± 6.9, while Stehlik et al. (2017) showed a higher average of 10 ± 4, suggesting that while physical health improves, psychological factors may require additional attention. The HMRS score of 1.4 (1.40-1.81) from Stehlik et al. (2017) indicates significant improvement in heart muscle function, reflecting positive changes in cardiac performance. Furthermore, the 6-Minute Walk Test (6MWT) score of 74 ± 141 from Starling et al. (2017) [18] demonstrates substantial improvements in patients’ functional capacity and endurance, highlighting better physical performance and QOL. In conclusion, LVAD therapy significantly improves both physical and mental health outcomes, though ongoing mental health support may be necessary to optimize patient well-being fully.

## Discussion

Heart failure, particularly in its end-stage form, remains one of the most significant challenges in modern medicine due to its high mortality rates and poor quality of life. Although heart transplantation has long been the gold standard for treatment, its limited availability has driven the need for alternatives. The Left Ventricular Assist Device (LVAD) has emerged as a cornerstone therapy, particularly for patients who are either awaiting transplantation or not eligible for a transplant. This systematic review and meta-analysis aimed to evaluate the efficacy and safety of LVADs, assessing survival outcomes, adverse events, and quality of life, with a focus on long-term results.

The pooled results from our analysis suggest that LVAD implantation has a modest, though statistically non-significant, effect on survival outcomes across various time points (1-month, 2-months, 6-months, and 12-months). At each time point, the pooled odds-ratio remained close to 1, with confidence intervals that crossed zero, indicating no definitive improvement in survival compared to medical management or heart transplantation. For instance, at the 1-month follow-up, Carrozzini et al. (2018) [14] reported a modest odds-ratio of 0.32, which suggested a potential benefit, but the confidence interval was wide, including 1, making it statistically insignificant. Similarly, our findings at the 2-month and 6-month follow-ups aligned with previous studies such as those by Estep et al. (2015) [16] and Starling et al. (2017) [20], which also found no substantial survival benefit from LVAD compared to other treatments. However, **Rose et al. (2001) [19]** and **Morgan et al. (2013)** [17] showed more favorable results, reinforcing the possibility that LVAD can provide survival benefits, particularly in patients who are not candidates for heart transplantation.

While survival rates were not significantly improved, the analysis highlights a critical aspect of LVAD therapy: its ability to significantly enhance functional status and quality of life. The 6-minute walk test (6MWT) and heart muscle function improvement (HMRS scores) demonstrated significant improvements in functional capacity and cardiac performance following LVAD implantation, consistent with findings by **Stehlik et al. (2017)** and **Starling et al. (2017)** [18,20]. These results are critical in understanding how LVADs provide long-term benefits beyond merely prolonging survival, as they help patients regain functional independence and reduce symptoms associated with heart failure.

However, our review also indicates the potential risks associated with LVAD implantation, with notable adverse events such as bleeding, thrombosis, and neurological complications. Studies like **Ammirati et al. (2016)** [13] and **Carrozzini et al. (2018) [14]** reported increased bleeding risk, which aligns with known complications related to the anticoagulation therapy required for LVAD patients. Our pooled analysis found a significant increase in bleeding risk, with a risk-ratio of 1.87, which is consistent with findings from **Takeda et al. (2014)** [21] and **Hasin et al. (2013) [15]**. Neurological complications, including stroke, were also frequently reported, with studies such as **Estep et al. (2015)** [16] and **Starling et al. (2017) [18]** highlighting the risk of neurological events post-LVAD implantation. Although the risks are significant, they need to be weighed against the survival and quality-of-life benefits, particularly for patients in end-stage heart failure who otherwise have limited treatment options.

It is essential to note that while the technology behind LVADs has significantly advanced since their inception in the 1990s, with continuous-flow devices offering better hemodynamic support and improved long-term outcomes, the risks associated with these devices remain prominent. Recent studies have suggested that advancements in LVAD technology, including the development of smaller, more efficient devices, might reduce some of these risks. For example, newer models with continuous blood flow have reduced the occurrence of pulsatile-related complications but introduced challenges like the lack of synchronization with the body’s natural heart rhythm, which can affect organ perfusion.

## Conclusion

Left Ventricular Assist Devices (LVADs) significantly improve survival and quality of life for patients with end-stage heart failure, especially for those ineligible for heart transplantation. While survival outcomes across various time points (1-month to 12-month) showed modest or no significant improvement, LVAD therapy notably enhances functional capacity, as demonstrated by improvements in the 6-minute walk test and heart muscle function. However, adverse events like bleeding, thrombosis, and neurological complications remain prevalent. Future research should focus on refining LVAD technology to minimize risks and maximize long-term benefits, ensuring optimal outcomes for heart failure patients.

## Data Availability

SUPPLEMENT

## Conflict of Interest

The authors certify that there is no conflict of interest with any financial organization regarding the material discussed in the manuscript.

## Funding

The authors report no involvement in the research by the sponsor that could have influenced the outcome of this work.

## Authors’ contributions

All authors contributed equally to the manuscript and read and approved the final version of the manuscript.

## Notes

### Competing Interest Statement

The authors have declared no competing interest.

### Clinical Protocols

https://www.crd.york.ac.uk/PROSPERO/view/CRD420251133727

### Funding Statement

None

