## supplementary file for "Impact of Left Ventricular Assist Devices on Quality of Life in End-Stage Heart Failure Patients: A Comprehensive Assessment"

Search String

("Left Ventricular Assist Device" OR "LVAD" OR "Mechanical Circulatory Support") AND ("End-stage Heart Failure" OR "Severe Heart Failure" OR "Advanced Heart Failure") AND ("Survival" OR "Quality of Life" OR "Adverse Events" OR "Complications").

Table S1. Summary and Demographic Characteristics

| **Author** | **Country** | **Study Type** | **Comparison** | **LVAD Used** | **Design of LVAD** | **Total Population** | **Treatment Comparison Males** | **Treatment Comparison Females** | **Average LVEF** | **Average Age** | **Main Finding** |
| --- | --- | --- | --- | --- | --- | --- | --- | --- | --- | --- | --- |
| Ammirati et. Al. 2015 | Italy | Observational Study | HTx | CF - LVAD | BTT | 213 | 44 | 112 | 24 ± 5 | 51 ± 5 | The study found that there was no statistically significant difference in mid-term survival between patients treated with continuous-flow LVAD and those who underwent heart transplantation (HTx). LVAD therapy provided comparable survival outcomes despite worse preoperative conditions, and it remains a valid option due to the scarcity of heart donors. |
| Ammirati et. Al. 2016 | Italy | Observational Study | HTx | CF-LVAD | BTT | 301 | 88 | 26 | 23 ± 3 | 57 ± 7 | The study found that CF-LVAD with BTT indication resulted in significantly better 1- and 2-year survival rates compared to heart transplantation using donors >55 years. However, LVAD patients experienced higher rates of rehospitalization and infection. |
| Carrozzini et. Al. 2018 | Italy | Observational Study | Medical Management or HTx | CF-LVAD | BTT | 126 | 47 | 64 | 27 ± 4 | 55 ± 4 | The study found that the use of new-generation intrapericardial continuous-flow left ventricular assist devices (CF-LVAD) as a bridge-to-transplant resulted in satisfactory outcomes both pre- and post-heart transplant (HTx). The survival rate on the waiting list at 6 months was 91%, and post-HTx survival at 1 year was 88%. |
| Hasin et. Al. 2013 | United States | Observational Study | Medical Management or HTx | Heartmate II | BTT | 115 | 96 | 96 | 17 ± 8 | 62 ± 7 | The study found that readmission rates for LVAD patients decreased significantly in the first 6 months following implantation and then stabilized. Leading causes of readmission were bleeding (66 readmissions), cardiac-related (51 readmissions), and infections (32 readmissions). There were also thrombotic events, including pump-related thrombosis |
| Estep et. Al. 2015 | United States | Observational Study | Medical Management or HTx | Heartmate II | BTT | 200 | 75 | 71 | 25 ± 7 | 64 ± 8 | LVAD therapy showed better survival and functional improvement, along with significant quality of life and depression reductions compared to OMM. However, it was associated with more frequent adverse events, particularly bleeding and neurological issues. |
| Morgan et. Al. 2013 | United States | Observational Study | Medical Management or HTx | CF-LVAD | BTT | 105 | 78 | 78 | 33.1 ± 4.9 | 53.8 ± 11 | The study found significant improvements in right ventricular function following CF- LVAD implantation, with reductions in CVP, PAP, and RVEDD, and increases in RVEF and RVSWI. Survival was better for patients without RV failure, but no specific adverse event data were provided. |
| Starling et. Al. 2017 | United States | Observational Study | Medical Management or HTx | Heartmate II | BTT | 200 | 97 | 103 | 27 ± 5 | Older | The main finding of the study is that LVAD therapy significantly improved survival and functional status compared to optimal medical management (OMM) at 2 years, with 70% survival in the LVAD group versus 41% survival in the OMM group (p < 0.001). Additionally, 30% of LVAD patients met the primary endpoint of survival with improvement in 6-minute walk distance (≥75 meters) compared to only 12% of OMM patients (odds ratio: 3.2, p = 0.012). |
| Rose et. Al. 2001 | United States | Randomnised Control Trials | Medical Management or HTx | Heartmate | BTT | 129 | 50 | 11 | 17 ± 7 | 68 ± 8.2 | The trial showed a 48% reduction in the risk of death in the LVAD group compared to the medical therapy group. At 1-year follow-up, survival was 52% in the LVAD group and 25% in the medical therapy group, with a significant improvement in quality of life for the LVAD group. |
| Stehlik et. Al. 2017 | United States | Observational Study | Medical Management or HTx | Heartmate II | BTT | 200 | 75 | 71 | N/A | 63 ± 13 | The main finding of the ROADMAP study is that LVAD therapy significantly improved health-related quality of life (hrQoL) in patients with low baseline self-reported QoL (VAS <55), but did not show a benefit for patients with higher baseline QoL (VAS ≥55). Survival outcomes at 12 months were similar between LVAD and optimal medical management (OMM) groups. |
| Takeda et. Al. 2014 | United States | Observational Study | Medical Management or HTx | CF-LVAD | BTT | 140 | N/A | N/A | 15.9 ± 6 | 54.7 ± 14.4 | The study found that continuous-flow LVAD therapy provided long-term survival (83% at 1 year, 75% at 3 years) but was associated with frequent rehospitalizations and adverse events like bleeding and stroke. Despite these challenges, renal and hepatic function improved during therapy |
| Stone et. Al. 2024 | Multiple Countries | Randomnised Control Trials | Placebo Procedure | CF-LVAD | BTT | 508 | 162 | 157 | 45.4 ± 8 | 74 ± 7 | The study found that transcatheter implantation of the Ventura interatrial shunt was safe but did not improve clinical outcomes in patients with heart failure overall. However, it reduced cardiovascular events in patients with reduced LVEF and increased mortality and hospitalizations in those with preserved LVEF. |

Table S2. Summary Findings of odds ratio after 1 month of LVAD implantation

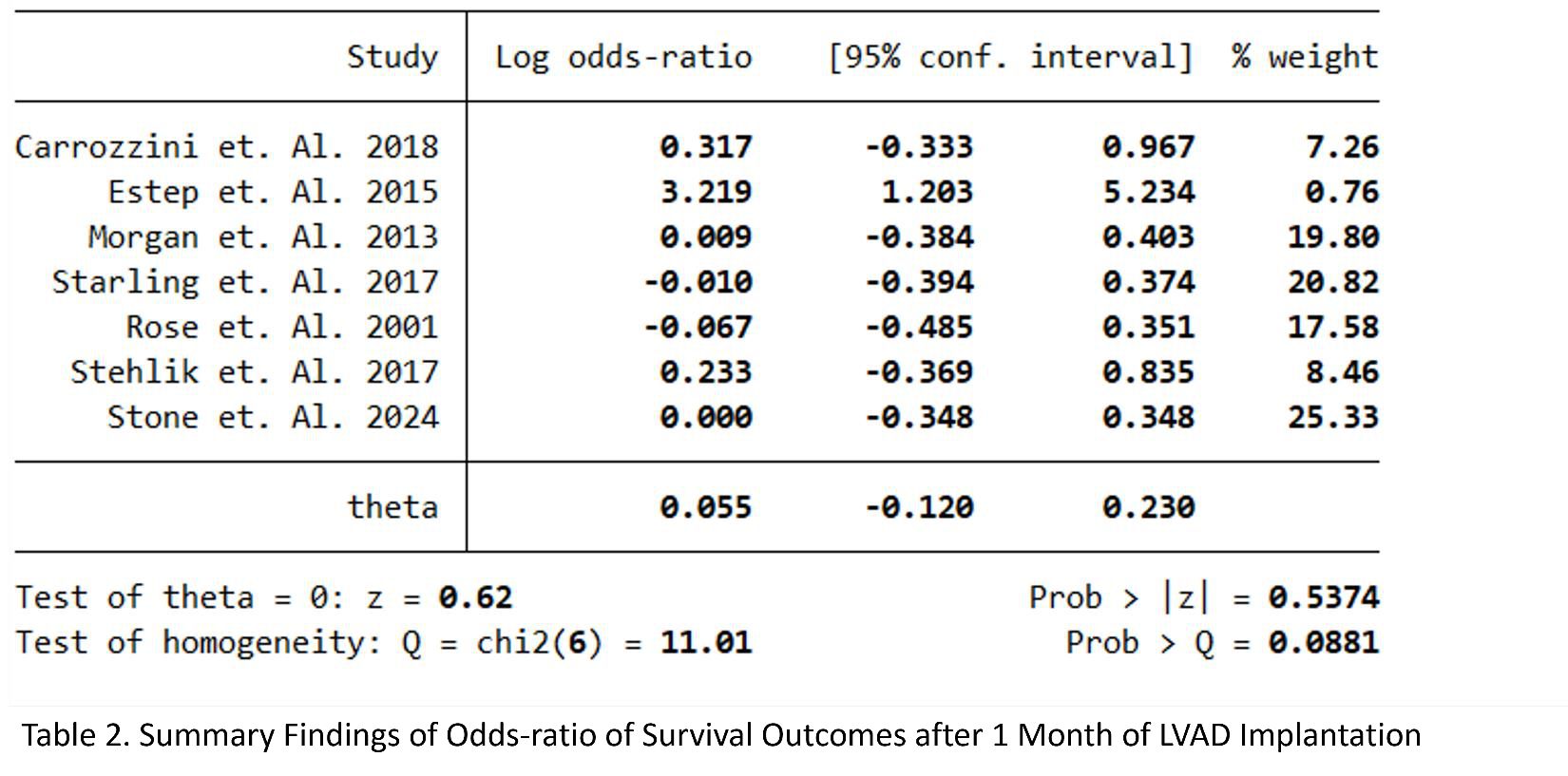

Table S3. Summary table of odds ratio of survival outcomes after 2 month of LVAD Implantation

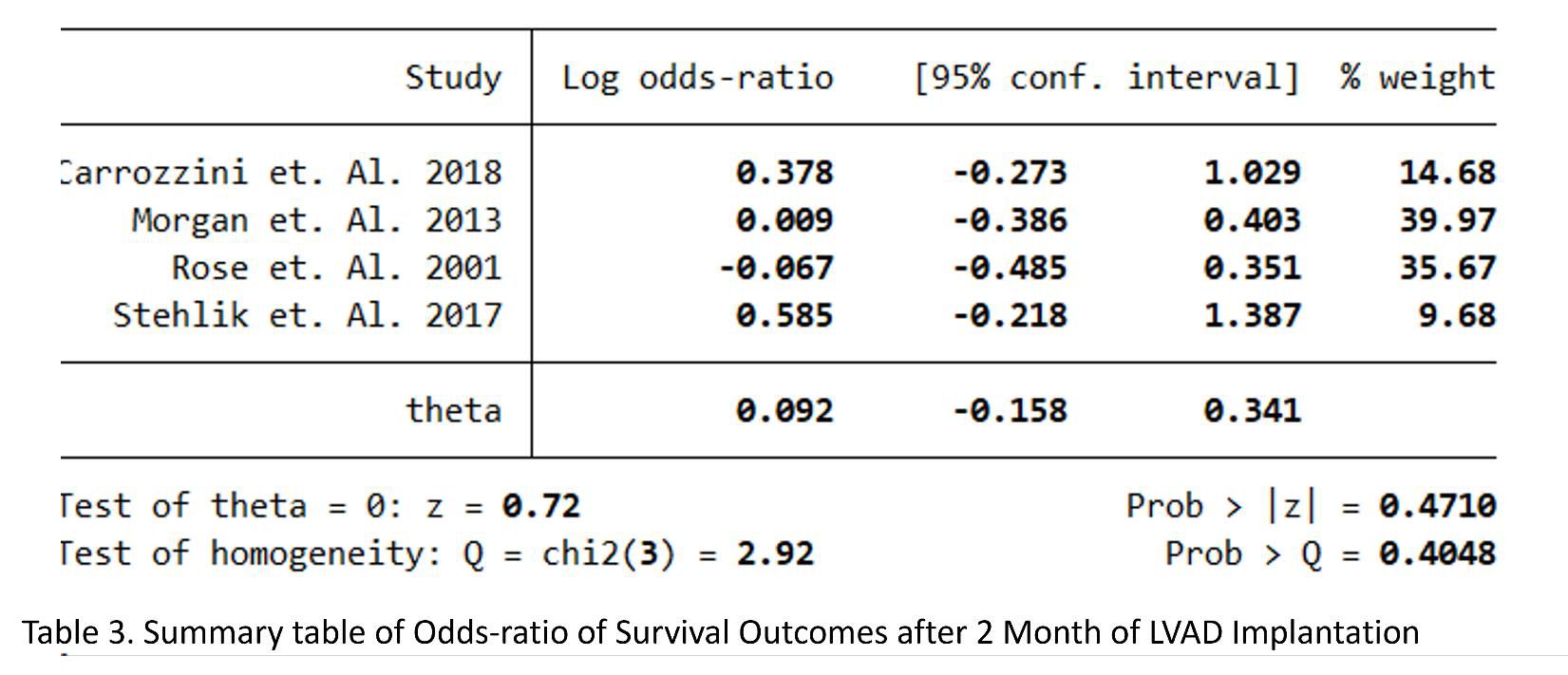

Table S4. Summary table of odds ratio of survival outcomes after 6 months of LVAD implantation

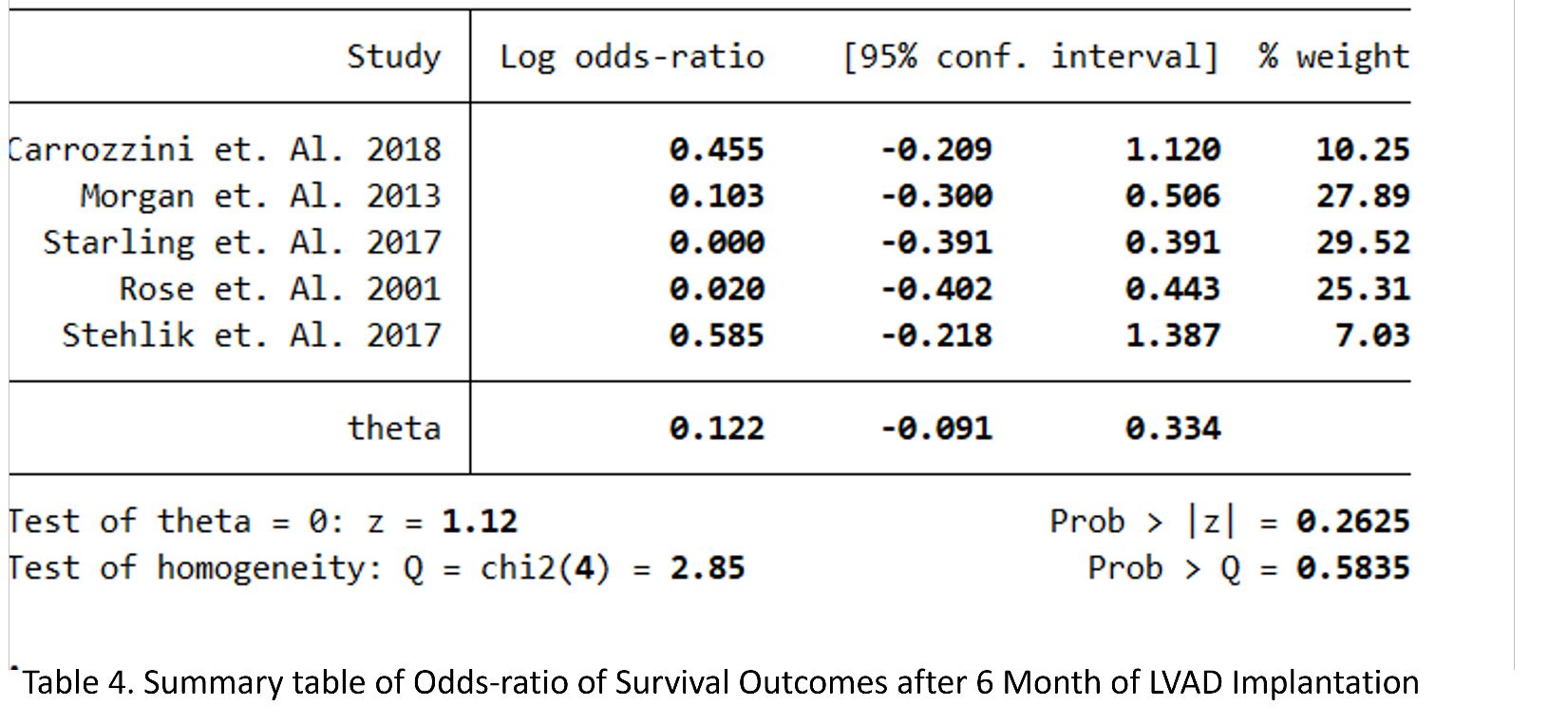

Table S5. Summary table of odds ratio of survival outcomes after 12 month of LVAD implantation

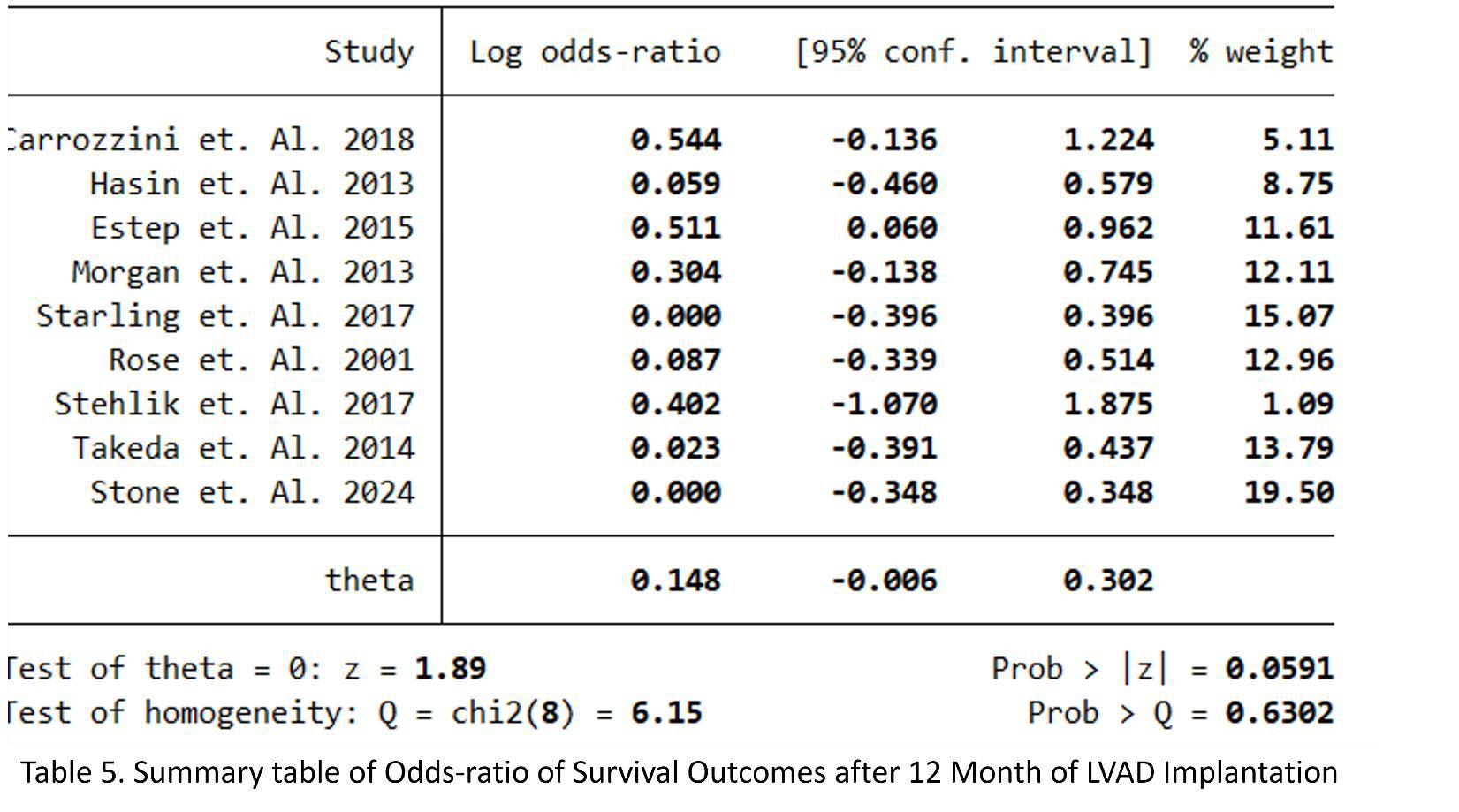

Table S6. Summary table of Odds risk ratio of death as adverse events in LVAD implantation

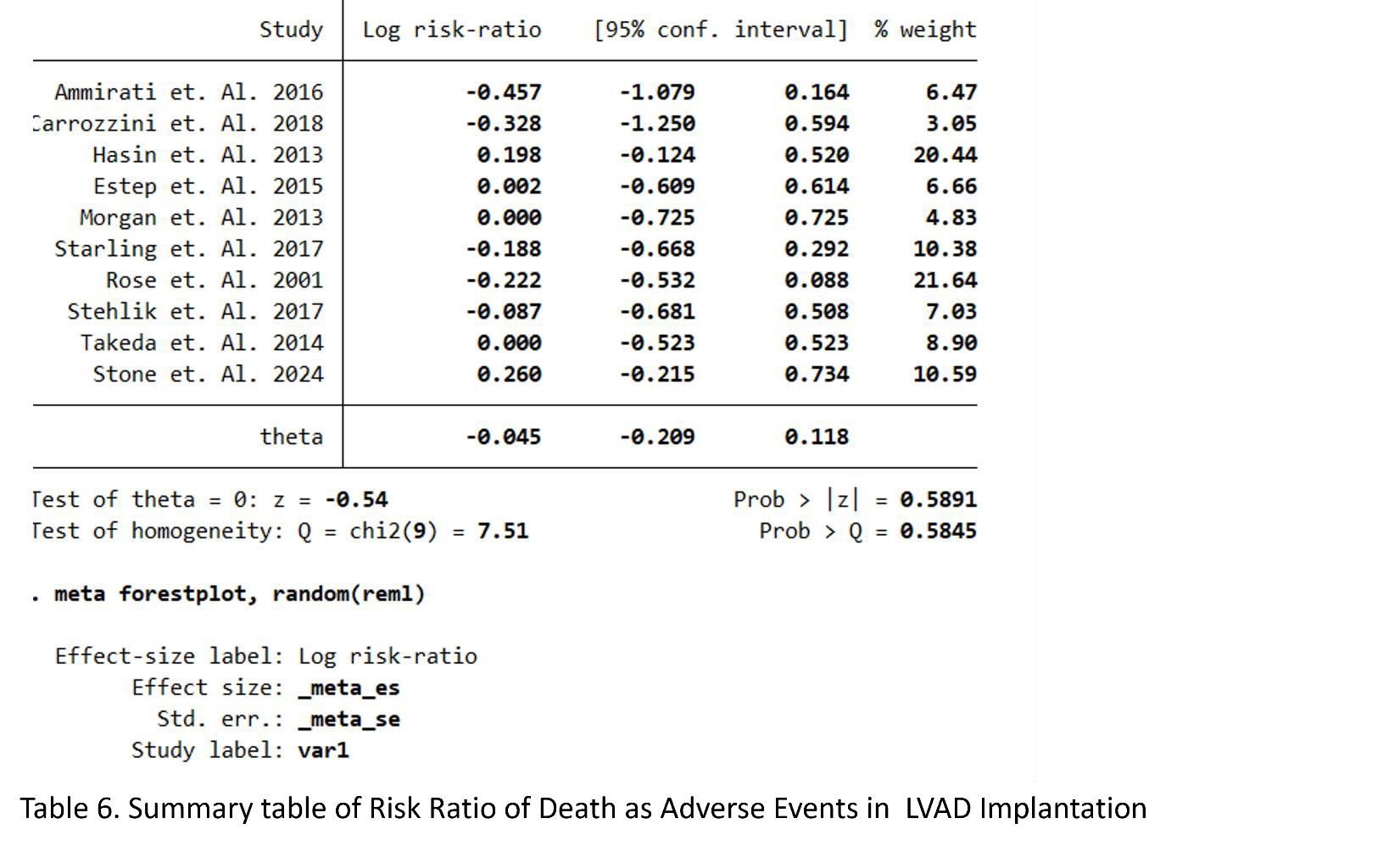

Table S7. Summary Table of Risk Ratio of Thrombotic as Adverse events in LVAD Implantation

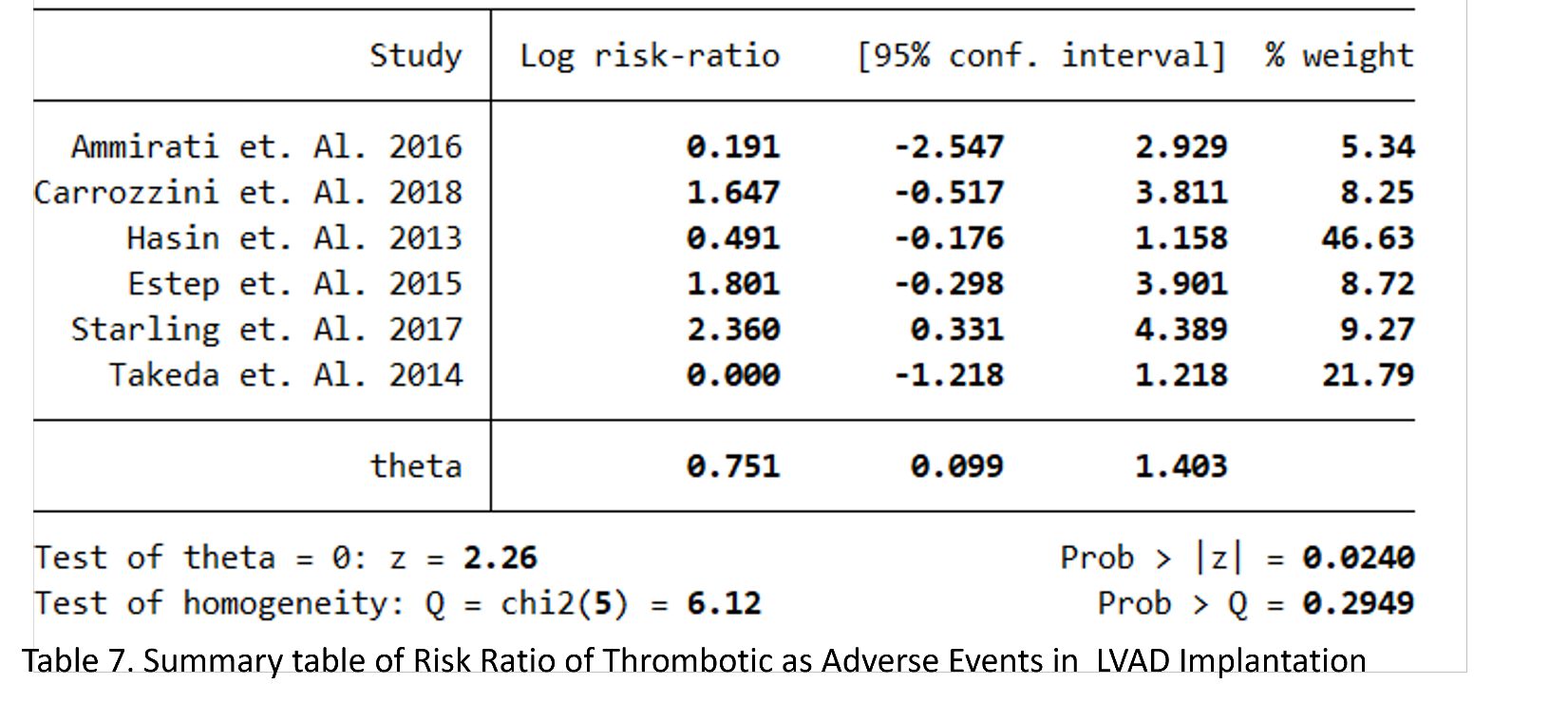

Table S8. Summary table of risk ratio of neurological consequences as adverse events in LVAD implantation

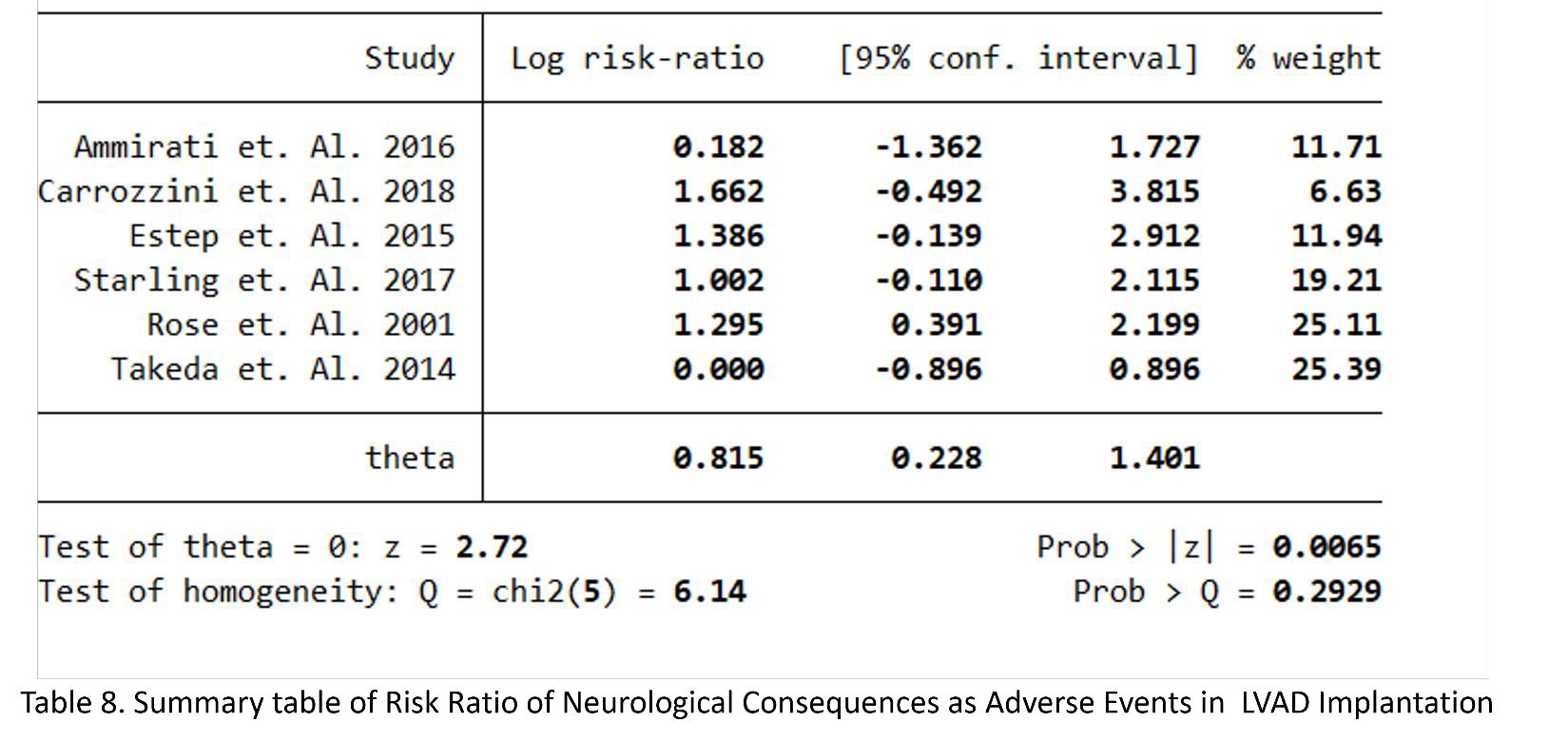

Figure S1 Risk of Bias with ROBINS-I

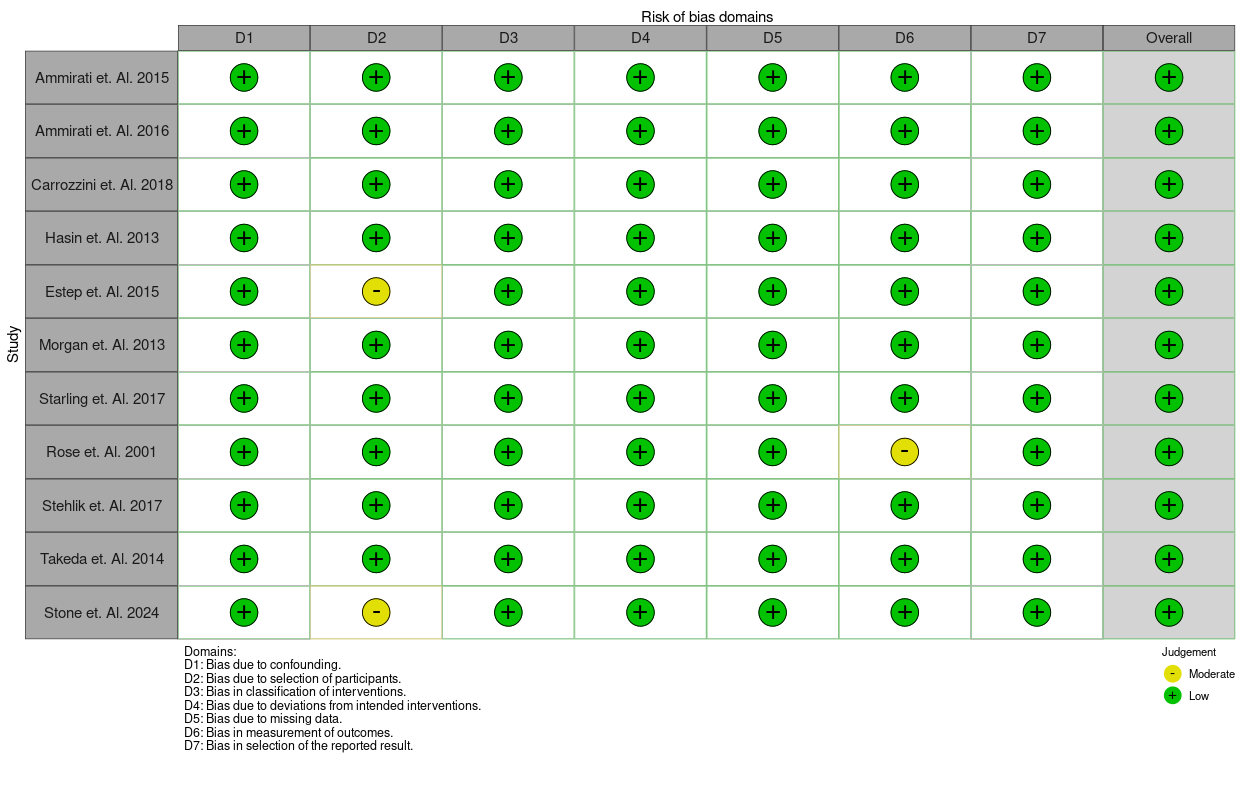
